# Wearable sensing for quantifying cognitive and balance functions in naturalistic movements of older adults with mild cognitive impairment in therapeutic environments

**DOI:** 10.64898/2026.06.18.26355980

**Authors:** Jangwon Lim, Rahul Islam, Dharini Raghavan, Bolaji Omofojoye, Amy D. Rodriguez, Yashar Kiarashi, Rachel Hershenberg, Gari D. Clifford, Hyeokhyen Kwon

**Author notes:** These authors contributed equally to this work (Jangwon Lim, Rahul Islam).

## Abstract

Mild cognitive impairment (MCI) is a clinically important stage preceding Alzheimer’s disease and related dementias, in which cognitive and balance functions are commonly evaluated using standard clinical assessments such as the Montreal Cognitive Assessment (MoCA) and Mini-Balance Evaluation Systems Test (Mini-BESTest). These assessments are administered episodically by clinicians and may miss functional changes during everyday movement. Recent studies and prior work in the Charlie and Harriet Shaffer Cognitive Empowerment Program (CEP), a therapeutic environment supporting lifestyle intervention and naturalistic social interaction, suggest that wearable and passive behavioral sensing can monitor movement patterns associated with cognitive and balance function in older adults with MCI. However, it remains unclear whether passive waist-mounted IMU data collected during naturalistic movement and social interaction can quantify clinician-rated cognitive and balance outcomes, particularly at the subdomain level, in an interpretable and demographically fair manner. To address this gap, we analyzed weekly IMU recordings collected over 6 months from 44 older adults with MCI in the CEP and trained tree-based ensemble regression models to estimate MoCA and Mini-BESTest total and subdomain scores, with interpretability and demographic fairness evaluation. Our models achieved RMSEs of 3.677 for MoCA and 3.672 for Mini-BESTest, benchmarked against Minimal Detectable Change and Minimal Clinically Important Difference thresholds. Feature importance analysis showed distinct movement signal properties across assessments, with general movement intensity features most informative for MoCA and temporal gait features led by cadence most informative for Mini-BESTest. Demographic bias analysis identified sex-related model bias, mitigated through post-processing while maintaining performance. This study supports the feasibility of wearable-based estimation of clinical assessment scores in older adults with MCI during naturalistic activity, with comparable performance between sexes after bias mitigation. This advances the validation of passive sensing for home monitoring to support clinical decision-making and personalized interventions.

**Author summary:** Mild cognitive impairment (MCI) is an early stage of cognitive decline that may precede Alzheimer’s disease and related dementias. Cognitive and balance changes are usually evaluated during clinical visits, but clinical assessments may miss functional changes that occur during everyday movement. In this study, we examined whether waist-worn motion sensors could be used to estimate standard cognitive and balance assessment scores in older adults with MCI during naturalistic activity in a therapeutic program. We used machine learning models to analyze movement patterns collected over six months and evaluated whether the models could estimate total and subdomain scores from the Montreal Cognitive Assessment and Mini-Balance Evaluation Systems Test. We also examined which movement features were most informative, whether estimations differed by sex or race, and whether post-processing bias mitigation reduced these differences. Our findings suggest that passive wearable sensing may provide useful information about cognitive and balance function of older adults with MCI in naturalistic settings. With further validation, this approach could support future monitoring and triage tools by helping identify individuals with MCI who exhibit concerning movement or balance changes and may benefit from more detailed clinical evaluation.

## Introduction

Alzheimer’s disease and related dementias (ADRD) represent an escalating global health crisis. As of 2021, more than 55 million people worldwide live with dementia [1, 2]. Mild cognitive impairment (MCI) marks the clinically identifiable phase preceding ADRD [3], with individuals converting to Alzheimer’s disease (AD) at an estimated annual rate of 10–15% per year [4]. MCI is screened by clinician-administered cognitive assessments, such as the Montreal Cognitive Assessment (MoCA) [5]. Because individuals with MCI often show gait and balance impairments [6, 7], clinician-administered balance assessments, such as the Mini-Balance Evaluation Systems Test (Mini-BESTest) [8], are widely used to evaluate these associated mobility and postural control deficits. Therefore, monitoring changes in cognitive and balance function within the MCI may help identify a critical window for lifestyle interventions and emerging therapies that may slow the progression of cognitive decline [9, 10]. However, these assessments require clinician administration, often not available for individuals living in regions with a limited number of specialists [11, 12], and occur episodically, which may miss subtle, day-to-day fluctuations in cognitive function, mobility, and postural control [13, 14].

To bridge the gap between episodic clinical visits and limited monitoring of functioning in daily life, wearable and artificial intelligence (AI) / machine learning (ML)-based monitoring systems have emerged as a promising alternative, which allow for the continuous, unobtrusive, and passive monitoring of behavioral markers in naturalistic settings [15]. Gait is a complex motor activity requiring attention, memory, and executive control [16, 17], and even subtle changes, like slower walking speed or increased stride-time variability, correlate with cognitive decline in MCI [18]. Wearable inertial measurement units (IMUs) have been shown to be effective in capturing such gait markers [19, 20] that are influenced by neural and cognitive function in real-world settings [18, 21], which may serve as a complement to episodic clinical visits. Yet several critical gaps remain. First, most prior studies analyzing behavioral data have focused on group-level classification tasks (e.g., MCI vs. healthy or higher-vs. lower-functioning MCI cohorts) [17, 22, 23] rather than quantifying clinician-rated scores like the MoCA or Mini-BESTest within individuals with MCI. This classification framing does not support tracking subtle changes in cognitive and balance function over time, which is central to clinical decision-making and quality of life in MCI. Second, prior wearable studies have been limited to controlled tasks administered by clinicians in laboratory settings [17, 22, 24], leaving their effectiveness in “living-lab” environments underexplored. Third, while some gait parameters have been linked to cognitive decline [6, 25–27], to our knowledge, prior work has not studied the association with or estimation of the subdomains of cognitive function defined in MoCA (e.g., Memory, Attention, Visuospatial, Executive, Language, and Orientation) or balance function defined in Mini-BESTest (e.g. Anticipatory, Sensory Orientation, Dynamic Gait, and Postural Control). Finally, existing studies rarely report and address biases in wearable-or passive-sensing AI models, concerning demographic variables (e.g., sex, race), which is critical for ensuring equitable deployment in diverse populations [28, 29].

To address these challenges, this study investigates two research questions: (RQ1) Can waist-mounted IMU data collected during naturalistic movements and social interactions be effectively used to quantify MoCA and Mini-BESTest and their subdomains? and (RQ2) To what extent do these predictive models demonstrate interpretability through digital biomarkers and ensure equitable performance across diverse demographic subgroups (sex and race)? To answer RQ1–2, we present a wearable sensing and evaluation framework studied with the waist-mounted IMU data collected from older adults with MCI for a 6-month period in a therapeutic space spanning 1700*m*^2^, where our participants can freely move and socially interact throughout the day outside of therapeutic programs. To the best of our knowledge, this study represents the first work on evaluating interpretable and demographically fair wearable-sensing models for quantifying cognitive and balance function in individuals with MCI during naturalistic movements.

## Related Works

### Associations of Gait and Balance with Cognitive Impairment in MCI

Motor and gait changes have been consistently observed in individuals with MCI and represent measurable behavioral correlates of cognitive function in this population. Several studies highlight that MCI is associated with measurable impairments in motor function, particularly gait and balance. Systematic reviews and meta-analyses have demonstrated that specific gait parameters such as velocity, stride length, and stride time significantly differ between older adults with MCI and normal cognition (NC), with dual-task paradigms enhancing discriminative power [6]. For instance, counting tasks during walking revealed greater sensitivity than verbal fluency tasks in detecting MCI-related gait changes, suggesting that cognitive load amplifies underlying motor dysfunctions. Bibliometric analyses have mapped the global evolution of this research domain, showing increasing interdisciplinary interest and identifying “dual-task walking” and “digital biomarkers” as emerging frontiers [30]. This line of research also underscores a shift from descriptive assessments of gait to predictive frameworks that incorporate machine learning and multicenter collaborations for standardization. Cross-sectional case–control studies further revealed that spatiotemporal gait variables, including step length and gait speed, can differentiate dementia from NC groups, though their discriminative ability between MCI and NC remains less robust [7].

Clinical observational studies support these findings, reporting that gait speed and step cycle parameters progressively worsen across the continuum of cognitive impairment, from subjective cognitive decline to MCI to major neurocognitive disorder [27]. Integrative approaches combining dual-task gait with eye-tracking have shown promise, achieving high classification accuracy in distinguishing cognitive impairment, and even linking gait features to plasma biomarkers such as phosphorylated tau [31]. Additional work has highlighted the association between cognitive impairment and global gait indices such as gait speed and walk ratio, suggesting that gait speed may be a more reliable indicator of central gait control [32]. Principal component analyses of spatiotemporal gait characteristics have identified distinct components, including variability, pace/stability, rhythm, and asymmetry, that capture gait heterogeneity in MCI compared to cognitively unimpaired individuals [25]. Such decomposition highlights step velocity variability, mean step length, and stance/swing asymmetry as potential core markers of MCI-related gait alterations. Beyond gait, studies also demonstrate that physical performance tests and balance assessments provide diagnostic and prognostic information for identifying early cognitive decline [33]. Importantly, evidence from systematic reviews suggests that physical activity and exercise may benefit cognitive and non-cognitive outcomes in individuals with MCI and dementia, further supporting the link between motor function and cognitive health [34]. A longitudinal study has examined the role of gait speed as a predictor of falls, finding that while older adults with MCI show slower gait compared to cognitively normal peers, short-term changes in gait speed alone may not reliably discriminate fallers within this group [26]. Taken together, the literature converges on the notion that MCI is accompanied by subtle but measurable changes in gait and balance, and digital markers derived from these signals offer a complementary modality for characterizing motor and cognitive function in this population. Previous research was mostly conducted in the context of dual-task assessments or controlled lab studies. This work extends understanding of the associations between gait and balance in MCI, in the context of naturalistic movements and social interactions, using wearable sensors, as a step toward evaluating the ecological validity of gait and balance assessment and monitoring in MCI populations.

### Detecting Cognitive Impairment and Balance Using Wearables and Passive Sensing with AI/ML Analysis

With advances in wearable sensors and time-series analysis techniques, many studies are exploring their use to quantify gait and balance associated with cognitive impairments in MCI. A systematic review of 49 studies on AI and wearable devices for early detection of cognitive impairment and dementia concluded that continuous passive monitoring shows significant promise for early risk stratification, yet highlighted critical methodological gaps including small sample sizes, short monitoring durations, and a lack of external validation [35]. A single lower-back inertial sensor was shown to capture gait features sensitive to cognitive decline, demonstrating the feasibility of wearable-derived digital biomarkers for characterizing cognitive impairment [36]. Building on this work, Jeon *et al.* [22] demonstrated that seven body-worn IMUs measured during straight-line, obstacle-negotiation, and various dual-task walking scenarios could detect early stages in AD through an ensemble machine learning analysis. A smart insole equipped with pressure sensors and IMUs was further used to classify the full spectrum of cognitive decline from cognitively normal to MCI to AD through deep learning, with classification accuracy increasing under more complex dual-task conditions [37]. For balance assessment, a deep learning model applied to smartphone-based inertial sensor data during the Mini-BESTest tasks showed the feasibility of classifying fall risk in older adults [38]. However, these studies remain confined to controlled laboratory or clinical settings.

While lab-based studies offer controlled conditions, they may not reflect the complexity and variability of real-world gait patterns, motivating the need for ecological validation of wearable-derived digital markers. In a free-living environment, wrist-worn accelerometer data combined with tree-based machine learning models achieved high discriminative power for predicting poor cognitive performance, particularly in processing speed and working memory [39]. A large-scale longitudinal cohort study using wrist-worn accelerometers further showed that free-living gait features, including slower maximal walking speed, increased step-time variability, and early bedtimes, were independently associated with incident dementia over a median follow-up of 7.5 years [40]. Xie *et al.* [18] also demonstrated that such gait differences could be captured using wearable sensors worn during daily outdoor walking over two weeks. While different from fully free-living monitoring, Hegde *et al.* [23] also evaluated privacy-preserving camera-based sensing in free movements and social interactions within therapeutic buildings spanning 1700*m*^2^ (the same study site as in this work), which was designed to provide lifestyle interventions resembling naturalistic settings, like kitchen, dining, library, etc. In this setting, camera-based sensing captured movement and social interaction patterns that distinguished higher-from lower-functioning MCI cohorts, demonstrating that passive behavioral sensing carries information about cognitive function. Together, these studies suggest that passive sensing approaches can capture clinically meaningful behavioral markers beyond controlled settings, although monitoring durations, sensing modalities, and analytic targets vary widely.

Beyond diagnostic group classification, wearable sensors have been used for continuous clinical assessment across a range of neurological and psychiatric conditions. Heart rate and accelerometer data have been combined to assess symptom severity in schizophrenia, enabling continuous symptom tracking over time [41]. Heart rate variability and body movement from a wearable chest patch have been used to estimate severity in Rett syndrome, showing that combined physiological and motor signals carry information about condition-specific functional status [42]. Wearable physiology and movement measured by accelerometers in real-world settings have also been analyzed to characterize challenging behaviors in individuals with autism spectrum disorder, demonstrating that multimodal wearable signals can capture clinically meaningful behavioral states [43]. In combination, these works establish wearable sensing as a viable approach for continuous, quantitative assessment across diverse neurological conditions, motivating its extension to cognitive and balance function in MCI.

Despite these advances, existing studies using passive sensing for MCI have predominantly focused on diagnostic (NC, MCI, or AD) or cohort-level classification rather than subject-level quantification of cognitive and balance profiles within MCI. This limits their utility for personalized interventions of MCI to slow down the cognitive decline. Equally important, wearable-based and passive sensing AI/ML systems need to be evaluated for their bias and fairness. Yuan *et al.* [28] demonstrated that ML models for AD progression exhibit significant fairness deficits across race and ethnicity, and broader evidence indicates similar issues across personal sensing devices [29]. Yet, assessing bias and studying its mitigation techniques within the fair ML framework remain largely unexplored for wearable-based quantification of gait, balance, and cognitive impairments in MCI. Together, these gaps point to the need for wearable and passive sensing approaches that move beyond condition classification, toward continuous quantification of cognitive and balance profiles with consideration of algorithmic fairness. Our work builds upon the current literature by deploying waist-mounted IMUs to capture naturalistic movements and social interactions in a therapeutic environment, quantifying continuous MoCA and Mini-BESTest scores including their domain-level subscores, with the goal of advancing ecological validity and fairness in wearable-based cognitive and balance assessment for MCI populations.

## Materials and methods

### Ethics Statement

This study recruited participants with MCI with approval from the Institutional Review Board (IRB) at Emory University (IRB #2025P010522) through a single IRB process. The participants provided their written informed consent to participate in this study. The participation period was between April 18th, 2024 and April 2nd, 2025.

### Dataset

#### Study Site and Participant Characteristics

This study was conducted in the Charlie and Harriet Shaffer Cognitive Empowerment Program (CEP) [44], a therapeutic program designed for individuals with MCI. Participants take part in structured weekly lifestyle intervention activities, tailored for MCI, including physical activity, cognitive training, functional independence training, nutrition education, and wellbeing education from 9:30 a.m. to 2:30 p.m. in a purpose-built 1700*m*^2^ indoor facility. The CEP program lasts for 6 months. Shown in Fig 1, the CEP includes a gym, kitchen, dining area, library, multipurpose rooms, resembling the spaces in daily living settings. The facility is instrumented with a distributed edge computing infrastructure [45] and indoor localization system [46] supporting multi-modal behavioral sensing, of which the present IMU-based study is one component. Therapeutic staff and caregivers are present throughout to provide support and ensure safety. The participants with MCI can freely interact and move around during lunch and break times outside of therapeutic programs on the day they visit the CEP. Participants were referred from the Cognitive Neurology Clinic in Emory Healthcare after receiving an MCI diagnosis based on Uniform Data Set (UDS) criteria from the National Alzheimer’s Coordinating Center. Cognitive and balance assessments were administered within CEP using the MoCA and Mini-BESTest at baseline (0 week) and at program exit (6 months). The CEP enrolls older adults diagnosed with MCI who vary in level of functioning, as assessed by neurologists in the Cognitive Neurology Clinic. We recruited 44 participants for this study.

**Fig 1.**
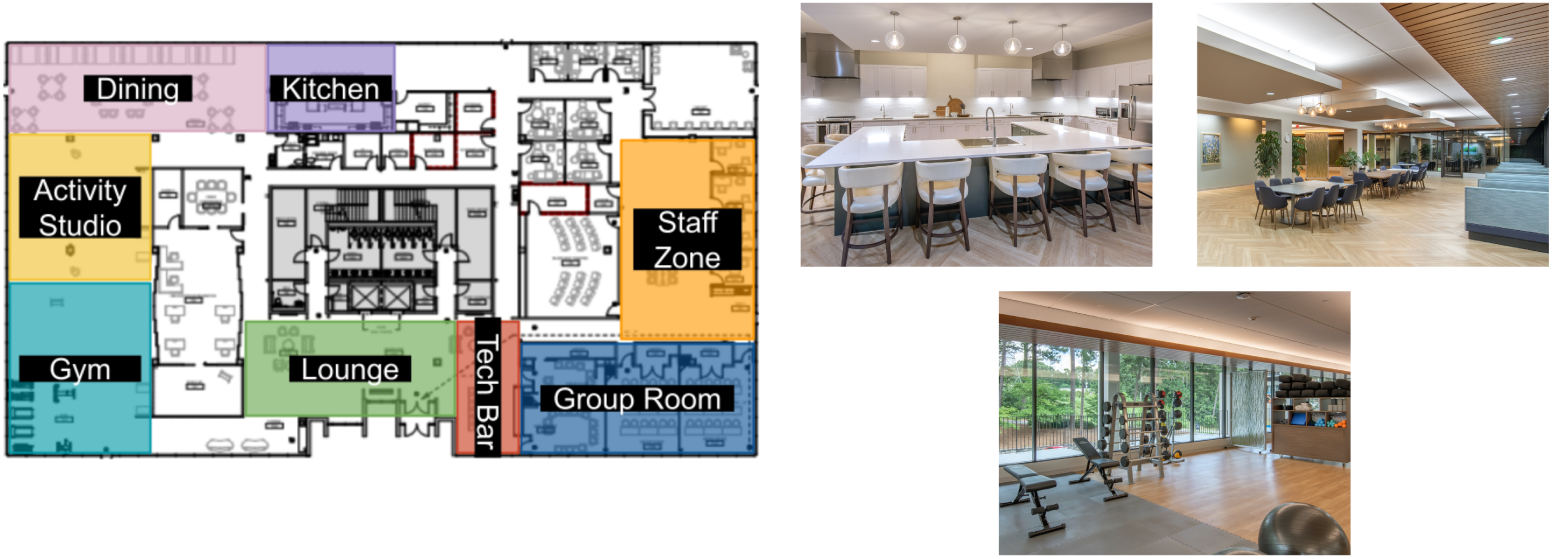
Spatial layout of the study space(CEP) and the exit total score was 21.3±4.1. Fig 2A shows the overall distribution of MoCA total scores at the baseline and exit for our participants. For Mini-BESTest (N=41), the baseline total score was 21.2±4.1, and the exit total score was 21.7±3.1. The subdomain scores of MoCA and Mini-BESTest are also reported in Table 1. Fig 2B shows the overall distribution of Mini-BESTest total scores at the baseline and exit for our participants.

**Fig 2.**
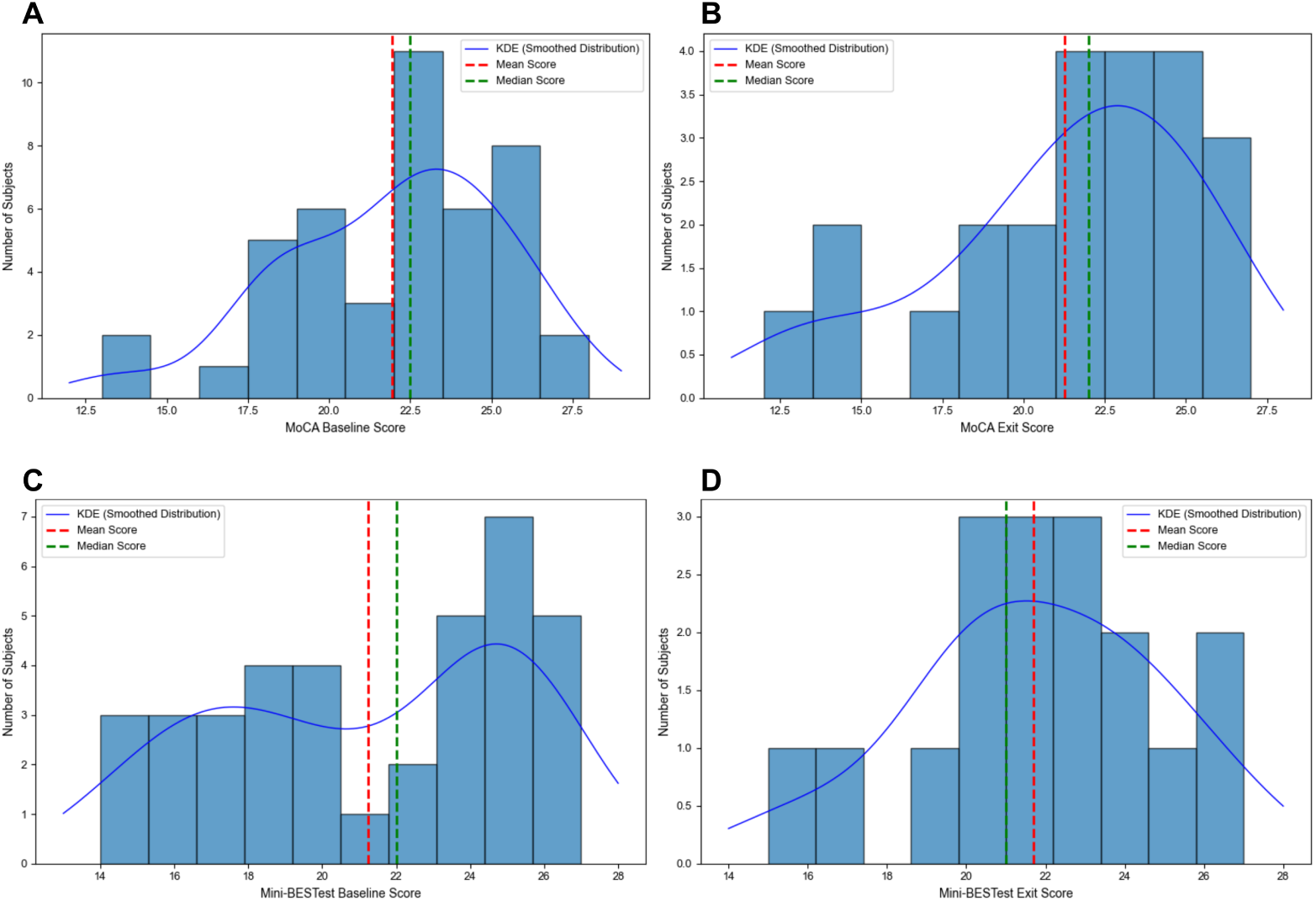
Total score distributions at baseline and exit. A: MoCA total score distribution at baseline. B: MoCA total score distribution at exit. C: Mini-BESTest total score distribution at baseline. D: Mini-BESTest total score distribution at exit.

**Table 1.** Demographic and clinical characteristics of the participants.

| Characteristic | MoCA (n=44) | Mini-BESTest (n=41) |
| --- | --- | --- |
| **Age (years), mean ± SD** | $74.1 \pm 6.6$ | $74.7 \pm 6.2$ |
| **Sex** |  |  |
| Male | 22 (50.0%) | 19 (46.3%) |
| Female | 22 (50.0%) | 22 (53.7%) |
| **Race** |  |  |
| White | 38 (86.4%) | 35 (85.4%) |
| Black / African American | 6 (13.6%) | 6 (14.6%) |
| **MoCA score, mean ± SD** |  |  |
| Baseline / exit total | $21.9 \pm 3.4$ / $21.3 \pm 4.1$ | — |
| Memory | $7.6 \pm 3.3$ | — |
| Executive | $10.5 \pm 2.3$ | — |
| Attention / concentration | $15.5 \pm 2.9$ | — |
| Language | $4.9 \pm 1.2$ | — |
| Visuospatial | $5.7 \pm 1.1$ | — |
| Orientation | $5.1 \pm 0.7$ | — |
| **Mini-BESTest score, mean ± SD** |  |  |
| Baseline / exit total | — | $21.2 \pm 4.1$ / $21.7 \pm 3.1$ |
| Anticipatory left, baseline / exit | — | $4.6 \pm 1.1$ / $4.6 \pm 1.1$ |
| Anticipatory right, baseline / exit | — | $4.5 \pm 1.1$ / $4.6 \pm 1.0$ |
| Reactive left, baseline / exit | — | $5.1 \pm 1.0$ / $5.0 \pm 1.2$ |
| Reactive right, baseline / exit | — | $5.0 \pm 1.2$ / $5.0 \pm 1.2$ |
| Sensory orientation, baseline / exit | — | $4.9 \pm 1.3$ / $5.4 \pm 0.9$ |
| Dynamic gait, baseline / exit | — | $6.9 \pm 1.8$ / $7.0 \pm 1.8$ |

#### Demographics, MoCA, and Mini-BESTest

The demographic and clinical characteristics, including MoCA and Mini-BESTest scores, are shown in Table 1. MoCA assesses multiple cognitive domains including memory([0, 15]), visuospatial ability([0, 7]), executive function([0, 13]), language([0, 6]), attention([0, 18]), and orientation([0, 6]) [47]. Scores below 26 indicate MCI, with lower scores reflecting increasing impairment severity [5]. For our participants, MoCA subdomain scores were available only at the exit assessment time point. Mini-BESTest evaluates dynamic balance across four systems: anticipatory postural adjustments([0, 6]), reactive postural control([0, 6]), sensory orientation([0, 6]), and dynamic gait([0, 10]) [48].

Shown in Table 1, 44 participants had MoCA scores assessed, and 41 participants were assessed with Mini-BESTest. For MoCA (N=44), the baseline total score was 21.9*±*3.4,

#### IMU Data Collection and Labeling

All participants visited the CEP weekly between 9am and 3pm. Upon arrival, the study team distributed waist-mounted IMU sensors (YOST Labs 3-Space Data Logger)^1^ in the morning and collected them before participants departed. The sensor was positioned near the lumbar region to approximate the body’s center of mass. The *Z*-axis was aligned vertically, with the anterior–posterior axis oriented forward during upright stance. To assess the ecological validity of quantifying MoCA and Mini-BESTest scores during free movement and social interactions, we used only 1 hour of IMU recordings captured during two scheduled breaks and lunchtime (10:55–11:10 and 12:00–12:45).

Because MoCA and Mini-BESTest assessments were administered at program entry and exit, we paired the IMU recordings from the first 3 months of each participant’s enrollment with their baseline scores and the recordings from the last 3 months with their exit scores, given their temporal proximity. This study design is intended to use the MoCA and Mini-BESTest scores as regression targets for IMU-derived features.

#### IMU Analysis Pipeline for Quantifying MoCA and Mini-BESTest scores

The overall analysis pipeline is shown in Fig 3. The raw IMU streams, including 3-axis accelerometer and 3-axis gyroscope (23 Hz), are preprocessed for noise removal, and segmented into windows with a mean duration of 3.41 seconds (SD = 1.36 s) and a mean overlap of 1.71 seconds (50%), considering sensor dropouts or rest periods. Each analysis window is labeled with the corresponding MoCA and Mini-BESTest total and subdomain scores (either baseline or exit, according to the time of recording). We extracted gait, statistical, and time-frequency features from each sensor window following previous work [39, 49, 50]. These features are processed with ML models (e.g., Random Forest, XGBoost) for regressing MoCA and Mini-BESTest scores. The window-level score regressions are aggregated across the entire recording for each subject, for subject-level evaluation of both baseline and exit scores. Our trained model was further assessed with SHAP-based feature importance analysis to identify the most informative features in free movements and social interactions associated with MoCA and Mini-BESTest scores, as well as the demographic biases (sex and race) of the model and bias mitigation analysis for developing a demographically fair model within a fair ML framework [28, 51].

**Fig 3.**
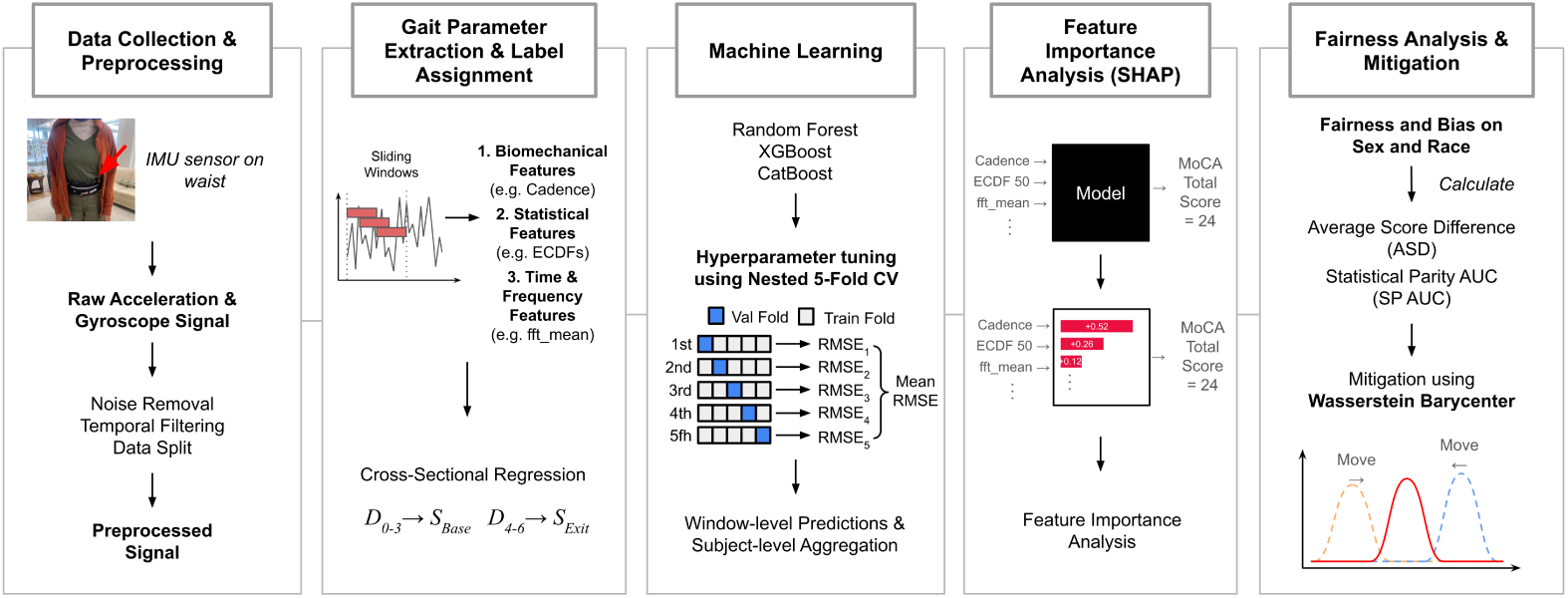
Overall data processing and analysis pipeline. The pipeline proceeds from raw IMU signal preprocessing and sliding-window feature extraction, where *D* represents datasets within a specific interval in months and *S* represents either baseline or exit scores. The subsequent stages involve machine learning model training using nested 5-fold cross-validation, interpretability analysis using SHAP values, and a fairness/bias evaluation and Wasserstein Barycenter mitigation to reduce demographic biases in model regressions.

#### Preprocessing

The 6-axis IMU time series was segmented into windows of 100 samples with 50% overlap. This window size was chosen to capture at least one stride (defined as one full gait cycle from one foot’s initial contact with the ground to the next contact of the same foot) within each window, as the average stride duration of older adults ranges from 1.08 s to 1.16 s depending on the gender [52]. To prevent the inclusion of discontinuous data caused by sensor dropouts or rest periods, we discarded any window containing a time gap exceeding 60 seconds. This segmentation yielded valid windows with a mean duration of 3.41 s (SD = 1.36 s). We then applied a two-stage filtering process to all three accelerometer axes. Previous work characterized the spectral content of trunk acceleration during human movement and reported that the energy band associated with daily activity begins at approximately 0.3 Hz, with the bulk of acceleration power at the upper body during walking falling between 0.8 and 5 Hz [53]. Guided by these bounds, we applied a high-pass filter at 0.3 Hz to remove the quasi-static gravitational (DC) component and a low-pass filter at 5.0 Hz to attenuate high-frequency sensor noise while preserving the dynamic acceleration content relevant to body movement. For the gyroscope axes, only the 5.0 Hz low-pass filter was applied, as the gyroscope signal does not contain a gravitational DC component. Both filters were implemented as 4th-order zero-phase Butterworth filters, which involve applying the filter in both forward and reverse directions to eliminate phase distortion and prevent temporal shifts in the signal [54, 55]. Feature extraction primarily utilized the vertical axis (z-axis) of the accelerometer data, with spatial gait and movement quality features additionally incorporating the other accelerometer axes and tri-axial gyroscope, as detailed below.

#### Feature Extraction

We extracted a 40-dimensional feature vector from each window of a 3-axis accelerometer and a 3-axis gyroscope. Our features include gait, statistical/distributional, and time-frequency features, as shown in S1 Table.

#### Gait Features

We captured spatiotemporal characteristics in gait, such as rhythm, stability, and smoothness. They are divided into three main categories: temporal features (relating to the timing of the gait cycle), spatiotemporal features (combining time and distance), and other kinematic features (describing the motion’s quality and orientation). Temporal gait features were extracted using gait events identified from the filtered z-axis acceleration (vertical direction) via a local maxima detection algorithm (find peaks function, SciPy Signal Processing Library) [56]. Given the target population of older adults with MCI, who characteristically exhibit reduced gait speed [27], we used a minimum inter-peak distance of around 0.5 seconds (11 samples at 23 Hz), adapting the temporal constraint approach in Del Din *et al.* [57] to minimize false-positive detections from intra-step artifacts while remaining sensitive to the step rates expected in this clinical cohort. From the detected array of peaks, we extracted *Cadence (steps/min), Stride Duration (s), Stride Variability,* and *Gait Symmetry Index (GSI)* [58]. *Stance Time* and *Swing Time* were not directly measured but were derived from the average stride duration based on the established biphasic model of a normal gait cycle, which consists of approximately 60% stance phase and 40% swing phase [59].

Spatial gait features, specifically *Stride Length (m)* and *Gait Velocity (m/s)*, were estimated separately using the inverted pendulum model described in Zijlstra and Hof [49], which posits that the body’s Center of Mass (CoM) follows a circular arc during the single stance phase. Under this model, step length is estimated as 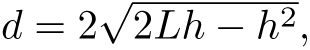 where L is the estimated leg length and *h* is the vertical displacement of the CoM during a single step cycle. Initial contact (IC) events were detected from the anterior-posterior (Y-axis) acceleration, low-pass filtered at 2 Hz using a fourth-order zero-phase Butterworth filter, by identifying zero-crossings from positive to negative values, which correspond to the transition from forward propulsion to braking at heel strike, and selecting the peak immediately preceding each zero-crossing as the IC event. We obtained the vertical displacement by double-integrating the low-pass filtered z-axis acceleration, followed by a high-pass filter at 0.1 Hz to remove integration drift. For each IC-to-IC step cycle, *h* was computed as the difference between the maximum and minimum displacement values. Because subject-specific anthropometrics were unavailable, leg length *L* was approximated from sex-and age-stratified population upper-leg lengths reported for adults aged 70–79 years (with total leg length estimated as twice the mean upper-leg length) [60], yielding *L* = 0.788 m for males and *L* = 0.708 m for females. Stride length was obtained by summing two consecutive step lengths, and gait velocity was computed as the ratio of mean stride length to mean stride duration, both derived from the same IC timestamps.

We further computed kinematic features to capture the quality and complexity of motion. We quantified movement smoothness by calculating the *RMS of the Jerk*, a proxy for gait stability [61]. In addition to linear motion from accelerometers, we also computed variables that represent rotational movement by incorporating the filtered tri-axial gyroscope data. We derived *Direction Change*, which captures the rate of change in rotational speed, and *Shannon Entropy of Gyroscope Magnitude*, which quantifies the diversity of rotational movement intensities within each window.

## Statistical Features

Statistical features were computed from the filtered vertical acceleration signal to characterize signal properties including distributional shape, temporal variability, and trend. We characterized the distribution of vertical acceleration’s amplitude using higher-order statistical moments, including *Skewness* and *Kurtosis* [62], as well as *Interquartile Range* to capture amplitude diversity and spread. We further extracted *Linear Trend Slope* to capture systematic drift within each window. A complete list of statistical features is provided in S1 Table. While the statistical features summarize the signal’s distribution using moments (e.g., skewness, kurtosis), these metrics can be disproportionately influenced by the extreme outliers common in data collected in naturalistic environments [63]. To acquire a more robust and non-parametric characterization of the signal’s amplitude distribution, we extracted the Empirical Cumulative Distribution Function (ECDF) features, widely used in wearable-based human activity recognition research [64]. We extracted a set of seven key percentiles: the 0th (minimum), 5th, 25th (first quartile), 50th (median), 75th (third quartile), 95th, and 100th (maximum), along with the mean and standard deviation of these ECDF values.

### Time-frequency Features

Time-frequency features were computed from the filtered vertical acceleration signal to capture temporal dynamics, signal complexity, and spectral properties. The signal’s overall power and magnitude were quantified by its *Absolute Energy*, *RMS Vertical Acceleration*, and *Absolute Maximum.* To measure signal volatility and smoothness, we computed the *Absolute Sum of Changes*, *Mean Absolute Change*, and the *Mean of the Second Derivative*. We captured the signal’s dynamic complexity and temporal dependence through its *Lag-1 Autocorrelation*, *Binned Entropy* (a histogram based Shannon entropy), *the Number of Peaks*, and *the Longest Strike above Mean*. For frequency features, we extracted the mean and variance of the absolute FFT coefficients after applying Fast Fourier Transform (FFT) over the signal filtered to 0.3–5 Hz, which reflect the overall magnitude and spread of spectral energy within the gait-relevant frequency range rather than energy in specific sub-bands. While these FFT-based features do not directly represent detailed gait characteristics, they complement time-domain features by capturing differences in the distribution of energy within the filtered band, which may reflect variations in movement regularity associated with MCI [36]. We also computed *Spectral Entropy* using Welch’s periodogram [65], which quantifies the uniformity of the signal’s power distribution.

### Regression of Subject-level MoCA and Mini-BESTest Scores

The window-level features served as input to tree-based ensemble models, including Random Forest (RF) [66], Categorical Boosting (CatBoost) [67], and Extreme Gradient Boosting (XGBoost) [68], to regress the MoCA and Mini-BESTest total scores as well as their subdomain scores (14 regression targets in total). For each subject, we aggregated and averaged window-level regressed scores into a single subject-level regression. For each best-performing model per target score, the Bland-Altman plots and the Pearson Correlation plots were generated on validation subject sets.

### Experiments and Evaluation Protocols

#### MoCA and Mini-BESTest Regression

The regression was evaluated through 5 runs of subject-wise nested 5-fold cross-validation. We split 64%, 16%, and 20% of subjects into training, validation, and test sets, respectively. The hyperparameter tuning of our tree-based ensemble models was conducted through the Optuna [69] library, on the training and validation sets. The left-out test set was used for reporting the final regression performance using Root Mean Square Error (RMSE) with 95% confidence intervals (CIs) to quantify estimation uncertainty. We evaluated all 14 regression targets including MoCA total and six subdomain scores and Mini-BESTest total and six subdomain scores. Subdomain-level analysis was included to capture the heterogeneity of impairment profiles across individuals, as subjects with similar total scores may exhibit distinct patterns of domain-specific deficits. To assess clinical utility, we benchmarked model performance against the Minimal Detectable Change (MDC), which distinguishes true change from measurement error, and the Minimal Clinically Important Difference (MCID), which reflects the smallest change that is clinically meaningful. We adopt established thresholds for MoCA total score (MDC: 5.1; MCID: 1.0) [70] and Mini-BESTest total score (MDC: 3.4; MCID: 3.8) [71]. To further assess model performance, we generated Bland-Altman plots [72] to examine the mean difference between predicted and observed scores and establish the limits of agreement, and Pearson correlation plots to assess the linear association between them. Both analyses were conducted on the cross-validated subject-level regressions.

#### Feature Importance Analysis

Identifying the most influential features (or digital markers) for regressing each clinical score [73], we used SHapley Additive exPlanations (SHAP) [74]. This analysis was conducted on the best-performing model (e.g., XGBoost, Random Forest) from the MoCA and Mini-BESTest score regression analyses. To ensure that subjects with more walking windows did not disproportionately influence the global importance rankings, we computed mean absolute SHAP values per subject first, then averaged across subjects with equal weight. We report the top features for each target using SHAP summary bar plots, and use beeswarm plots to illustrate the directionality of feature-score relationships at the window level.

#### Demographic Bias Analysis and Mitigation

Beyond predictive accuracy, we conducted a bias analysis of the best-performing models for MoCA and Mini-BESTest total score regression concerning demographic variables [28]. We studied two sensitive attributes, shown in Table 1: Sex (defined as “Male” and “Female”) and Race (defined as “White” and “Non-White”). For this analysis, Male and White participants were designated as the reference (privileged) groups, while Female and Non-White participants were considered the interest (unprivileged) groups. Specifically, we assessed the biases in regression using two regression-adapted metrics from the statistical parity framework [75]. Statistical Parity AUC (SP AUC) measures the disparity in probability distributions between groups, with lower values indicating greater fairness. Average Score Difference (ASD) measures the difference in mean predicted scores between the interest and reference groups, ASD (*µ*_interest_ *µ*_reference_), where a value of zero indicates perfect fairness, while a negative value implies systematic underestimation of the interest group’s scores. To address any identified disparities, we explored a post-processing mitigation strategy using the Wasserstein Barycenter method [51]. This technique learns an optimal transformation to align the statistical distributions of the regressed scores from the different demographic groups. For each target and sensitive attribute, the Wasserstein Barycenter model was fit using the regressed scores generated from the training set and their corresponding group labels. This learned transformation was then applied to the regression of the test set, which was evaluated by comparing the changes in SP AUC, ASD, and RMSE before and after bias mitigation.

## Results

### MoCA and Mini-BESTest Regression

#### MoCA

Table 2 shows the regression performance (RMSE) of the three tree-based ensemble machine learning models (Random Forest, CatBoost, and XGBoost) for predicting the MoCA total score and its six clinical subdomains. For regressing the total score, the RF model achieved an RMSE of 3.677 (95% CI: [3.404, 3.949]). Similarly, for the memory, attention, executive, language, visuospatial, and orientation subdomains, the RF model achieved the lowest RMSE across all targets, with values of 3.364, 2.764, 2.090, 1.150, 1.088, and 0.610, respectively. For the total score, Bland-Altman analysis (Fig 4A) showed a mean bias of 0.03 points with 95% limits of agreement of [ −7.36, 7.42], and Pearson correlation between predicted and observed scores was *r* = −0.19 with *p* = 0.156 (Fig 5A). Full Bland-Altman and Pearson correlation plots for all MoCA subdomains are provided in S1 Table and S3 Fig.

**Fig 4.**
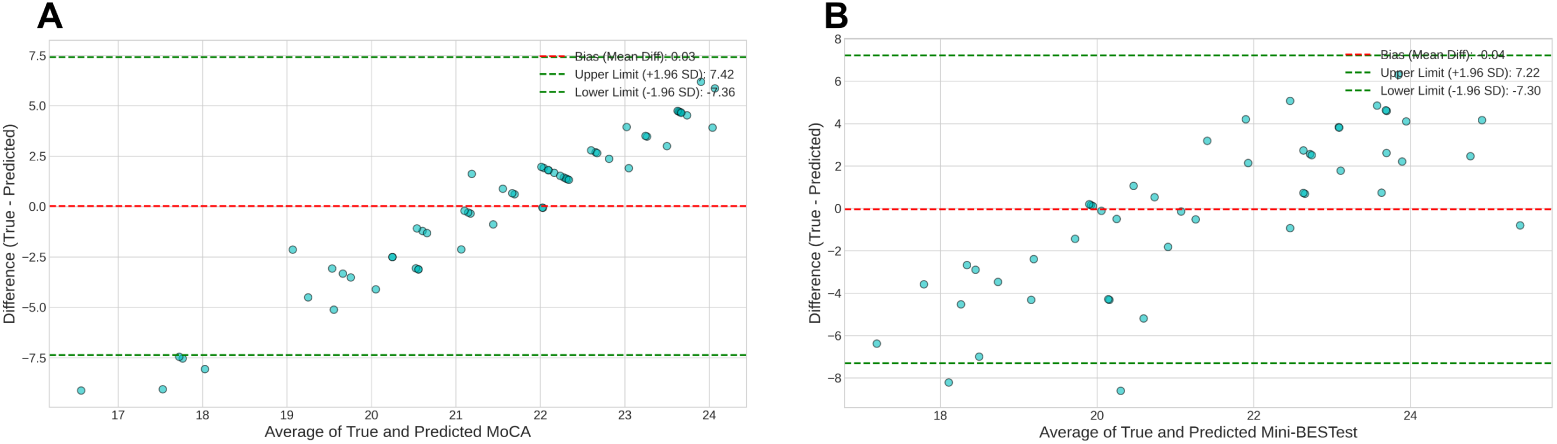
Bland–Altman plots for total score regression. A: Bland–Altman plot for MoCA total score regression using the random forest model. B: Bland–Altman plot for Mini-BESTest total score regression using the CatBoost model. The dashed lines indicate mean bias and 95% limits of agreement.

**Fig 5.**
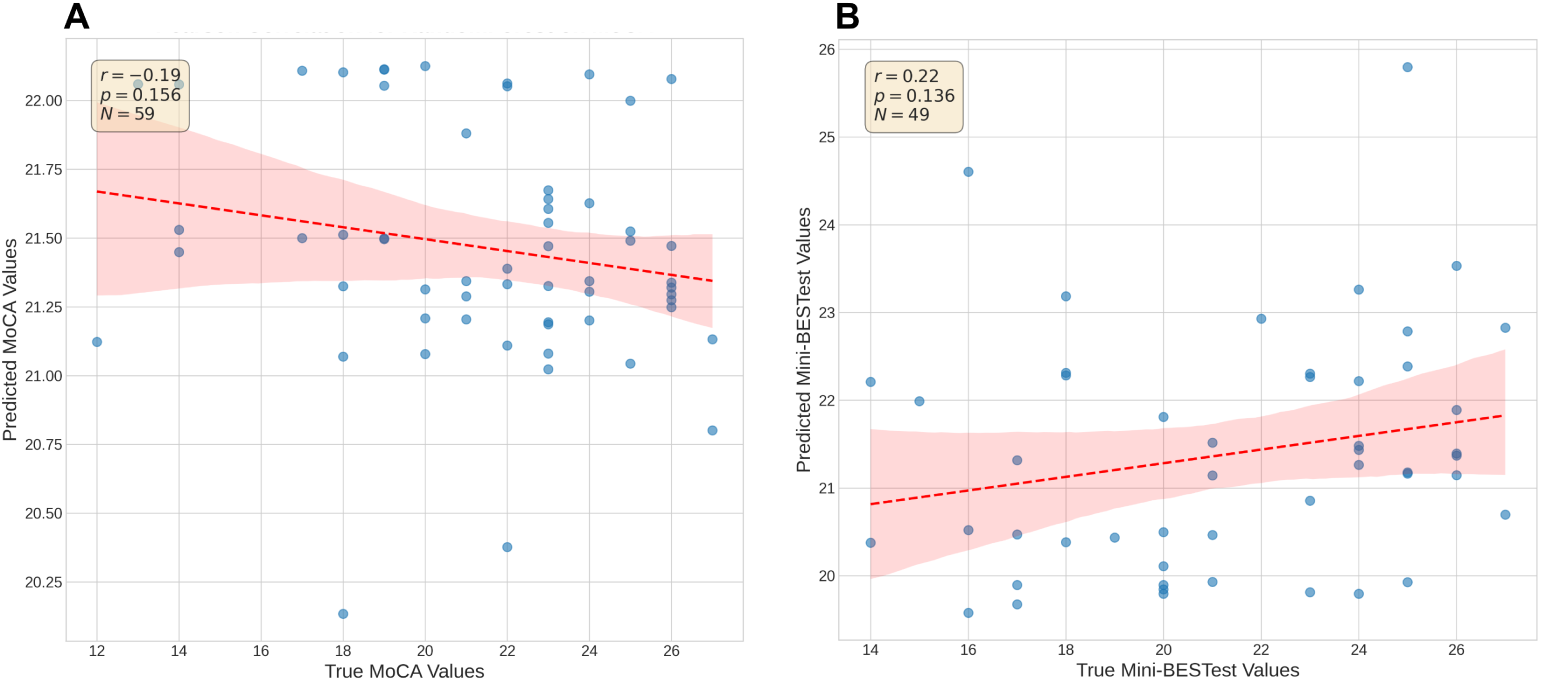
Pearson correlation between predicted and observed total scores. A: Pearson correlation for MoCA total score regression using the random forest model. B: Pearson correlation for Mini-BESTest total score regression using the CatBoost model. The regression line and 95% confidence band are shown for each assessment.

**Table 2.** Regression performance for MoCA and Mini-BESTest total and subdomain scores.

| Assessment | Score | Range | RF | CatBoost | XGBoost |
| --- | --- | --- | --- | --- | --- |
| MoCA | Total Score | [0, 30] | **3.677 [3.404, 3.949]** | 3.706 [3.445, 3.966] | 3.715 [3.450, 3.980] |
|  | Memory | [0, 15] | **3.364 [2.908, 3.820]** | 3.415 [2.938, 3.892] | 3.467 [2.972, 3.962] |
|  | Attention / Concentration | [0, 18] | **2.764 [2.346, 3.182]** | 2.932 [2.522, 3.342] | 2.927 [2.498, 3.355] |
|  | Executive | [0, 13] | **2.090 [1.722, 2.458]** | 2.216 [1.827, 2.604] | 2.202 [1.810, 2.594] |
|  | Language | [0, 6] | **1.150 [1.001, 1.300]** | 1.213 [1.069, 1.358] | 1.210 [1.062, 1.358] |
|  | Visuospatial | [0, 7] | **1.088 [0.961, 1.215]** | 1.140 [0.997, 1.283] | 1.138 [0.993, 1.282] |
|  | Orientation | [0, 6] | **0.610 [0.453, 0.767]** | 0.632 [0.508, 0.757] | 0.643 [0.520, 0.766] |
| Mini-BESTest | Total Score | [0, 28] | 3.750 [3.626, 3.873] | **3.672 [3.574, 3.771]** | 3.685 [3.595, 3.775] |
|  | Dynamic Gait | [0, 10] | **1.849 [1.779, 1.921]** | 1.866 [1.805, 1.927] | 1.866 [1.805, 1.928] |
|  | Sensory Orientation | [0, 6] | 1.332 [1.228, 1.437] | **1.311 [1.202, 1.420]** | 1.317 [1.208, 1.426] |
|  | Reactive Postural Control Right | [0, 6] | **1.061 [0.976, 1.146]** | 1.062 [0.980, 1.143] | 1.064 [0.984, 1.145] |
|  | Reactive Postural Control Left | [0, 6] | **0.983 [0.922, 1.043]** | 0.999 [0.939, 1.059] | 1.007 [0.948, 1.066] |
|  | Anticipatory Right | [0, 6] | 0.966 [0.885, 1.048] | 0.884 [0.804, 0.964] | **0.883 [0.804, 0.962]** |
|  | Anticipatory Left | [0, 6] | 1.042 [0.922, 1.163] | 0.979 [0.859, 1.099] | **0.975 [0.858, 1.092]** |

RMSE values are reported with 95% confidence intervals in brackets. Bold values indicate the best-performing model for each target.

### Mini-BESTest

Table 2 also presents the regression performance for predicting the Mini-BESTest total score and its six subdomain scores related to dynamic balance and gait. For the total score, the CatBoost model achieved the lowest RMSE of 3.672 (95% CI: [3.574, 3.771]). Among the subdomains, RF performed best on three targets, CatBoost on one (Sensory Orientation), and XGBoost on two (Anticipatory Right and Left). Subdomain RMSEs ranged from 0.883 (Anticipatory Right) to 1.849 (Dynamic Gait). For the total score, Bland-Altman analysis (Fig 4B) showed a mean bias of-0.04 points with 95% limits of agreement of [ 7.30, 7.22], and Pearson correlation between predicted and observed scores was *r* = 0.22 with *p* = 0.136 (Fig 5B). Full Bland-Altman and Pearson correlation plots for all Mini-BESTest subdomains are provided in S2 Fig and S4 Fig.

### Feature Importance Analysis

#### MoCA

Shown in Fig 6A, for the MoCA total score, statistical and time-frequency descriptors of the vertical acceleration signal were identified as the primary predictors. Absolute Sum of Changes recorded the highest importance score of 0.161, followed by Mean Absolute Change (0.133) and Autocorrelation Lag 1 (0.115). Fig 7A shows the SHAP beeswarm plots for each feature’s importance and their directionality. The SHAP analysis results for each MoCA subdomain are shown in S5 Fig and S6 Fig.

**Fig 6.**
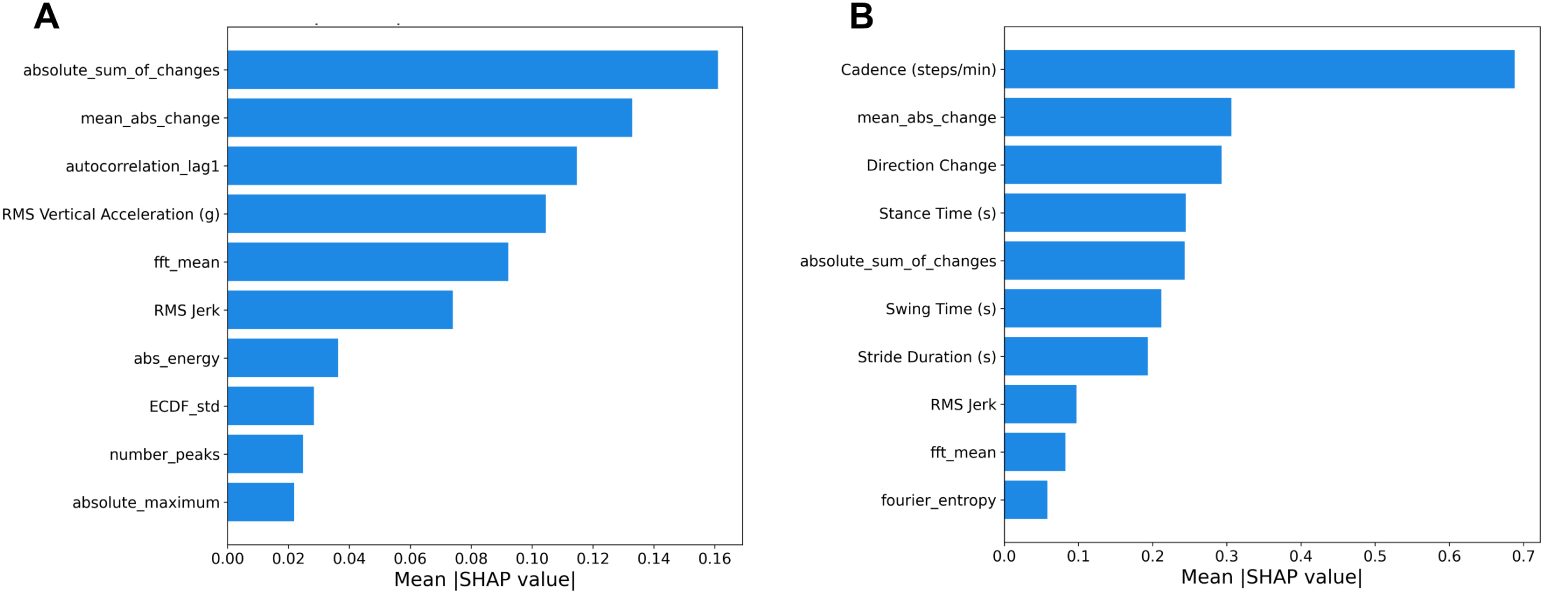
SHAP feature importance for total score regression. A: MoCA total score regression using the random forest model. B: Mini-BESTest total score regression using the CatBoost model. The plots show the top 10 features ranked by mean absolute SHAP value.

**Fig 7.**
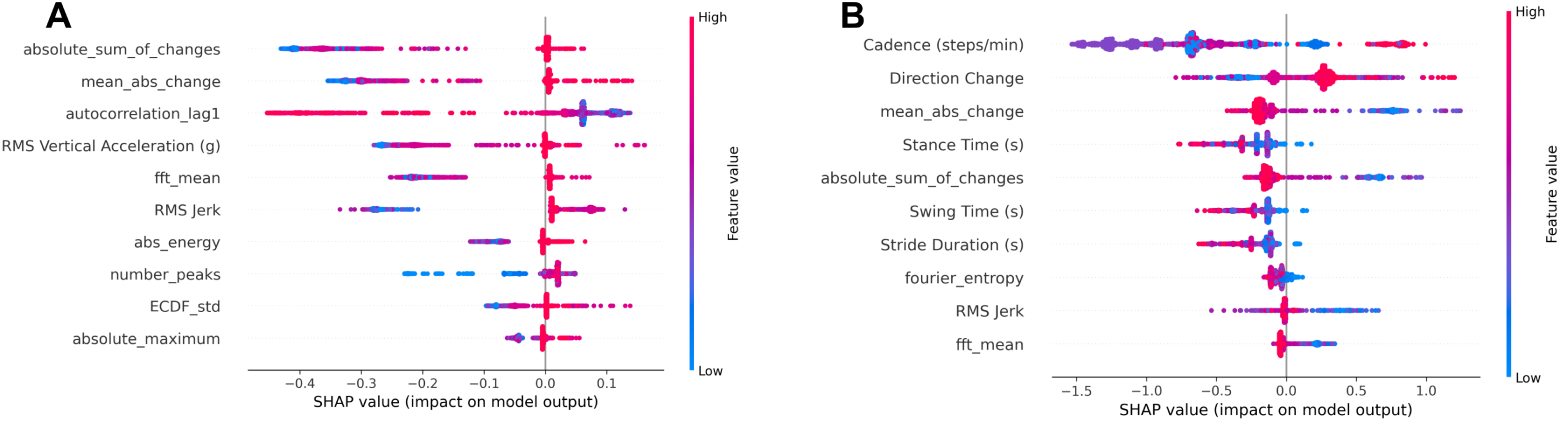
SHAP beeswarm plots for total score regression. A: MoCA total score regression using the random forest model. B: Mini-BESTest total score regression using the CatBoost model. The plots show the distribution and directionality of SHAP values for the top-ranked features.

### Mini-BESTest

The SHAP analysis result for the Mini-BESTest total score is shown in Fig 6B. Cadence (steps/min) was the primary predictor (importance score: 0.688), followed by Mean Absolute Change (0.306) and Direction Change (0.293). Fig 7B shows the SHAP beeswarm plots for each feature’s importance and their directionality relative to the Mini-BESTest total score. Feature importance plots for each Mini-BESTest subdomain are provided in S7 Fig and S8 Fig.

### Demographic Bias Analysis and Mitigation

#### MoCA

Table 3 shows the biases in the best regression models for the MoCA total score and subdomain scores across sex and race attributes, before and after bias mitigation. For sex, the ASD for MoCA total score regression before bias mitigation was 0.292 and moved toward zero after mitigation ( −0.010), with a corresponding SP AUC decrease from 0.143 to 0.033. RMSE remained nearly unchanged from 3.294 to 3.291. Similar reductions in SP AUC were observed across all the MoCA subdomain score regressions (e.g., Attention/Concentration: 0.206 to 0.091; Visuospatial: 0.232 to 0.087; Orientation: 0.209 to 0.092). ASD generally moved toward zero after mitigation (e.g., Attention/Concentration: 0.207 to 0.103; Language: 0.122 to 0.047; Visuospatial: 0.168 to 0.030), though some subdomains showed ASD crossing zero (e.g., Executive: 0.007 to 0.117; Memory: 0.066 to 0.081). For race, the ASD of MoCA total score regression was 0.003 and increased after mitigation to 0.172, while SP AUC decreased from 0.118 to 0.090 and RMSE decreased slightly (3.294 to 3.236). Several subdomain score regressions showed increased ASD after mitigation (e.g., Attention/Concentration: 0.037 to 0.274; Executive: 0.149 to 0.209; Language: 0.019 to 0.117), with mixed SP AUC changes (e.g., Memory SP AUC: 0.136 to 0.144; Attention/Concentration SP AUC: 0.111 to 0.122), while Visuospatial exhibited a notable SP AUC reduction (0.268 to 0.155). Orientation showed the smallest changes (ASD: 0.004 to 0.049; SP AUC: 0.181 to 0.146). RMSE differences before and after mitigation for both sex and race attributes were marginal.

**Table 3.**
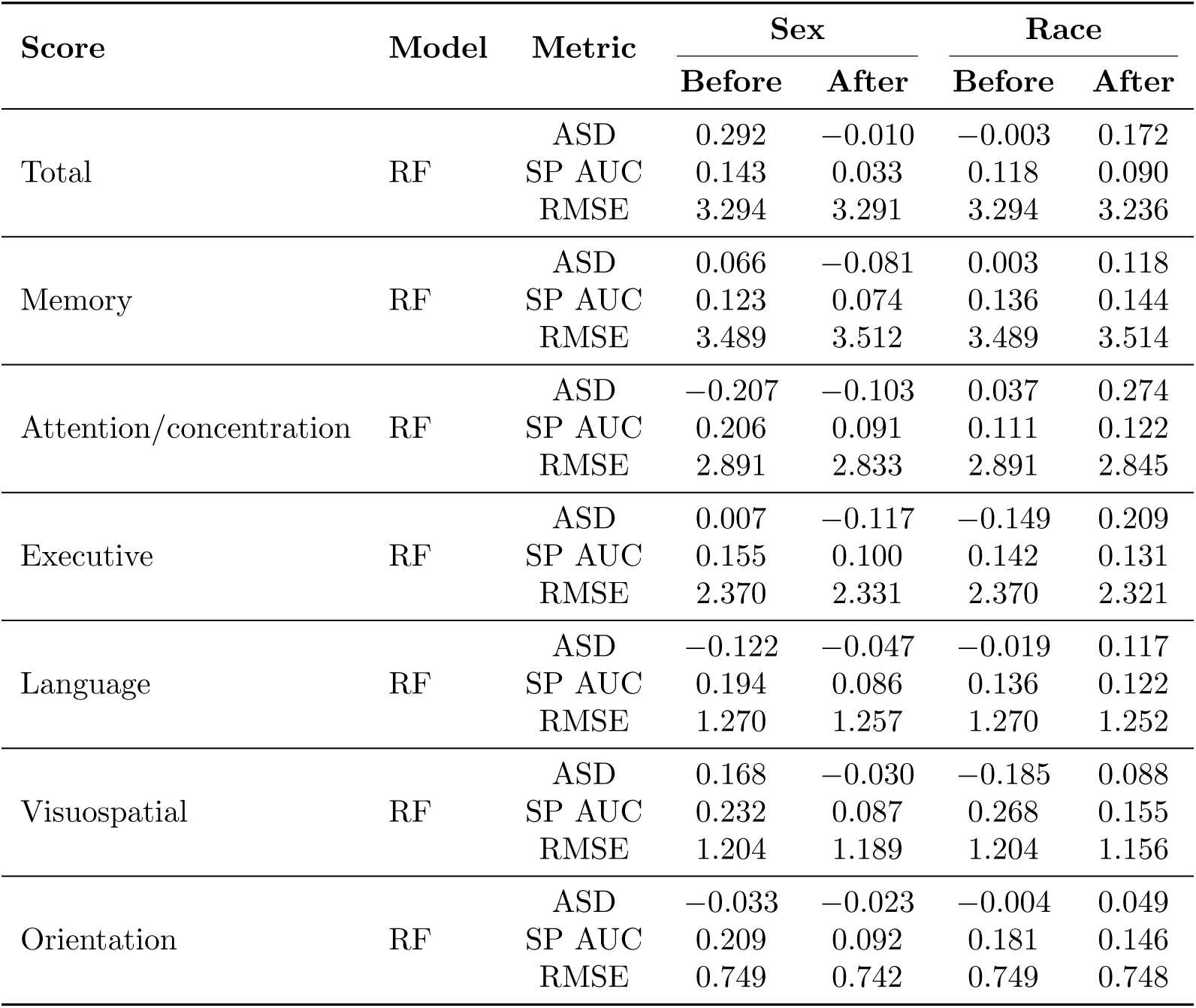
Bias analysis and mitigation results for MoCA total and subdomain scores. Values were computed us best regression model for each score.

| Score | Model | Metric | Sex |  | Race |  |
| --- | --- | --- | --- | --- | --- | --- |
|  |  |  | Before | After | Before | After |
| Total | RF | ASD | 0.292 | $-0.010$ | $-0.003$ | 0.172 |
|  |  | SP AUC | 0.143 | 0.033 | 0.118 | 0.090 |
|  |  | RMSE | 3.294 | 3.291 | 3.294 | 3.236 |
| Memory | RF | ASD | 0.066 | $-0.081$ | 0.003 | 0.118 |
|  |  | SP AUC | 0.123 | 0.074 | 0.136 | 0.144 |
|  |  | RMSE | 3.489 | 3.512 | 3.489 | 3.514 |
| Attention/concentration | RF | ASD | $-0.207$ | $-0.103$ | 0.037 | 0.274 |
|  |  | SP AUC | 0.206 | 0.091 | 0.111 | 0.122 |
|  |  | RMSE | 2.891 | 2.833 | 2.891 | 2.845 |
| Executive | RF | ASD | 0.007 | $-0.117$ | $-0.149$ | 0.209 |
|  |  | SP AUC | 0.155 | 0.100 | 0.142 | 0.131 |
|  |  | RMSE | 2.370 | 2.331 | 2.370 | 2.321 |
| Language | RF | ASD | $-0.122$ | $-0.047$ | $-0.019$ | 0.117 |
|  |  | SP AUC | 0.194 | 0.086 | 0.136 | 0.122 |
|  |  | RMSE | 1.270 | 1.257 | 1.270 | 1.252 |
| Visuospatial | RF | ASD | 0.168 | $-0.030$ | $-0.185$ | 0.088 |
|  |  | SP AUC | 0.232 | 0.087 | 0.268 | 0.155 |
|  |  | RMSE | 1.204 | 1.189 | 1.204 | 1.156 |
| Orientation | RF | ASD | $-0.033$ | $-0.023$ | $-0.004$ | 0.049 |
|  |  | SP AUC | 0.209 | 0.092 | 0.181 | 0.146 |
|  |  | RMSE | 0.749 | 0.742 | 0.749 | 0.748 |

#### Mini-BESTest

Table 4 shows the biases in the best regression models for the Mini-BESTest total and subdomain scores across sex and race attributes, before and after bias mitigation. For sex, the ASD for regressing Mini-BESTest total score before mitigation was 0.414 and moved toward zero after mitigation (−0.009), with SP AUC decreasing from 0.121 to 0.034. RMSE decreased slightly from 3.529 to 3.496. Reductions in sex SP AUC were consistently observed across all subdomains (e.g., Dynamic Gait: 0.212 to 0.041; Sensory Orientation: 0.145 to 0.040; Reactive Postural Control Right: 0.212 to 0.038), with ASD values moving toward zero in all cases (e.g., Dynamic Gait: 0.163 to 0.021; Sensory Orientation: 0.161 to 0.018; Anticipatory Right: 0.123 to 0.005).

**Table 4.** Bias analysis and mitigation results for Mini-BESTest total and subdomain scores. Values were comput the best regression model for each score.

| Score | Model | Metric | Sex |  | Race |  |
| --- | --- | --- | --- | --- | --- | --- |
|  |  |  | Before | After | Before | After |
| Total | CatBoost | ASD | $-0.414$ | $-0.009$ | $-0.439$ | $0.537$ |
| | | SP AUC | $0.121$ | $0.034$ | $0.175$ | $0.117$ |
| | | RMSE | $3.529$ | $3.496$ | $3.529$ | $3.481$ |
| Dynamic gait | RF | ASD | $-0.163$ | $-0.021$ | $0.005$ | $0.135$ |
| | | SP AUC | $0.212$ | $0.041$ | $0.128$ | $0.108$ |
| | | RMSE | $1.869$ | $1.884$ | $1.869$ | $1.870$ |
| Sensory orientation | CatBoost | ASD | $-0.161$ | $-0.018$ | $-0.244$ | $0.029$ |
| | | SP AUC | $0.145$ | $0.040$ | $0.209$ | $0.114$ |
| | | RMSE | $1.250$ | $1.242$ | $1.250$ | $1.247$ |
| Reactive postural control right | RF | ASD | $-0.144$ | $-0.006$ | $0.126$ | $0.014$ |
| | | SP AUC | $0.212$ | $0.038$ | $0.209$ | $0.118$ |
| | | RMSE | $1.074$ | $1.064$ | $1.074$ | $1.075$ |
| Reactive postural control left | RF | ASD | $-0.072$ | $-0.005$ | $0.189$ | $0.087$ |
| | | SP AUC | $0.107$ | $0.036$ | $0.356$ | $0.114$ |
| | | RMSE | $0.948$ | $0.946$ | $0.948$ | $0.936$ |
| Anticipatory right | XGB | ASD | $-0.123$ | $-0.005$ | $-0.166$ | $0.154$ |
| | | SP AUC | $0.110$ | $0.042$ | $0.135$ | $0.117$ |
| | | RMSE | $0.915$ | $0.892$ | $0.915$ | $0.943$ |
| Anticipatory left | XGB | ASD | $-0.018$ | $-0.004$ | $-0.049$ | $0.173$ |
| | | SP AUC | $0.080$ | $0.038$ | $0.098$ | $0.123$ |
| | | RMSE | $1.046$ | $1.053$ | $1.046$ | $1.054$ |

RMSE changes were marginal overall, with some subdomains showing slight decreases (e.g., Reactive Postural Control Right: 1.074 to 1.064; Sensory Orientation: 1.250 to 1.242) and others showing slight increases (e.g., Dynamic Gait: 1.869 to 1.884; Anticipatory Left: 1.046 to 1.053). For race, the ASD of Mini-BESTest total score regression was 0.439 before mitigation and changed direction after mitigation to 0.537, while SP AUC decreased from 0.175 to 0.117 and RMSE decreased from 3.529 to 3.481. Subdomain analysis showed mixed results. Some subdomains exhibited reduced SP AUC (e.g., Sensory Orientation: 0.209 to 0.114; Reactive Postural Control Left: 0.356 to 0.114), while others showed SP AUC increases (e.g., Anticipatory Left: 0.098 to 0.123). ASD after mitigation frequently crossed zero (e.g., Sensory Orientation: 0.244 to 0.029; Anticipatory Right: 0.166 to 0.154). RMSE differences before and after mitigation for race were marginal across all subdomains.

## Discussion

### MoCA and Mini-BESTest Regression

Our experiment demonstrates that ensemble tree-based regression models (Random Forest [66], CatBoost [67], XGBoost [68]) are highly effective in regressing both MoCA and Mini-BESTest scores. While all models exhibited comparable performance with marginal differences, RF and CatBoost achieved the lowest RMSE for the total scores.

#### MoCA

For regressing the MoCA total score, the RF model achieved the numerically lowest RMSE of 3.677 (95% CI: [3.404, 3.949]). This performance was statistically comparable to CatBoost (RMSE = 3.706) and XGBoost (RMSE = 3.715), suggesting that all three ensemble tree-based models captured the variance in overall cognitive scores with similar effectiveness. RF also consistently achieved the lowest RMSE across all six subdomains, with values ranging from 0.610 (Orientation) to 3.364 (Memory). This consistent advantage aligns with RF’s bagging-based regularization, which provides robust generalization under small-sample conditions [66] where gradient boosting methods are more susceptible to overfitting. Overall, the RMSEs on total and subdomain scores were below 4 points, with total score RMSEs below the MDC of 5.1 for the MoCA [70]. This level of error shows that passive IMU monitoring can approximate overall cognitive status for broad screening and stratification. However, the RMSE remains above the MCID of 1.0 [70] for MoCA total score, indicating that the model should not be interpreted as sensitive enough to reliably detect the smallest clinically meaningful changes or to replace comprehensive neuropsychological testing. Instead, the most defensible use case is triage, and wearable-derived estimates can serve as a continuous, low-burden monitoring signal that prompts in-depth clinical assessment upon potential changes in cognitive function larger than MDC.

MoCA subdomain analysis helps identify which components of the assessment are more or less inferable from passive gait-derived IMU features, but interpretation must explicitly account for differences in score range and score distribution. Several subdomain scores have small numeric ranges (e.g., Orientation [0–6], Language [0–6], Visuospatial [0–7]), and in our cohort some of these domains cluster near the upper end (e.g., Orientation at exit: 5.1±0.7; Visuospatial: 5.7±1.1; Language: 4.9±1.2). Under these conditions, a model can achieve a low RMSE partly by predicting the common near-ceiling value, so lower RMSE in these domains may not imply strong domain-specific sensitivity, which warrants further investigation with a larger sample size in the future. In contrast, domains with larger score ranges (e.g., Memory [0–15], Attention/Concentration [0–18], Executive function [0–13]) yielded RMSE values representing 22.4%, 15.4%, and 16.1% of their respective maximum scores, suggesting that naturalistic walking patterns may carry indirect signals related to memory, attention, and executive function [76, 77].

Bland-Altman analysis for the total score showed a near-zero mean bias of 0.03 points, indicating little average directional offset between predicted and observed scores. The 95% limits of agreement spanned [−7.36, 7.42], reflecting the variability inherent in estimating cognitive scores from passively collected movement data in a naturalistic setting with a limited sample size. The Pearson correlation between predicted and observed scores was not statistically significant (*r* =−0.19, *p* = 0.156), which is likely attributable to the restricted score range typical of a clinically homogeneous MCI cohort (*N* = 44). In such cohorts, score variability is compressed within a narrow band, which can attenuate correlation coefficients even when other performance metrics appear acceptable [72].

#### Mini-BESTest

For the Mini-BESTest total score, the CatBoost model achieved the lowest RMSE of 3.672 (95% CI: [3.574, 3.771]), outperforming Random Forest (RMSE = 3.750) and XGBoost (RMSE = 3.685). Although the confidence intervals overlapped, CatBoost demonstrated the lowest mean RMSE, suggesting its effectiveness in capturing the overall variance of mobility and balance functions. Unlike the MoCA results, where RF dominated all targets, the Mini-BESTest subdomains exhibited model variability. RF achieved the lowest RMSE for Dynamic Gait and both Reactive Postural Control tasks, XGBoost for both Anticipatory tasks, and CatBoost for Sensory Orientation. This diversity may reflect the heterogeneous nature of balance subdomains, where different movement patterns favor different model architectures. When benchmarked against established clinical thresholds, this error is slightly above the Mini-BESTest MDC of 3.4 but remains below the MCID of 3.8 [71]. This result suggests that the model may not yet be precise enough to reliably distinguish changes near the measurement-error threshold. However, because the RMSE is lower than the MCID, the model’s estimation error is smaller than the magnitude of change considered clinically meaningful. Therefore, the model is plausibly precise for detecting clinically meaningful differences in this MCI cohort.

Performance across the subdomains revealed the capacity of wearable sensing to capture distinct aspects of movement and balance. Models generally performed well on unilateral tasks, with RMSE values below or close to 1.0 for both Anticipatory (0.883 for Right, 0.975 for Left) and both Reactive Postural Control subdomains (1.061 for Right, 0.983 for Left). However, as with MoCA, the lowest RMSEs in these unilateral subdomain scores (RMSE ≈ 0.88–0.98 on 0–6 scales) must be interpreted with caution, as these subdomains had high average values in our cohort (e.g., exit reactive 5.0 1.2 out of 6 and exit anticipatory 4.6 1.1 out of 6). This is likely due to limited variability in our participants and needs further study with a larger sample size. In contrast, Dynamic Gait (RMSE = 1.849 on a 0–10 scale) showed relatively higher error, reflecting the inherent complexity and variability in quantifying dynamic walking patterns compared to isolated balance tasks, when using a single waist-mounted IMU sensor.

Similar to MoCA, Bland-Altman analysis for the Mini-BESTest total score yielded a mean bias of −0.04 points with 95% limits of agreement of [−7.30, 7.22]. This indicates that while the model introduces little average directional bias, the precision of any single estimate remains variable given the naturalistic setting and limited sample size. The Pearson correlation was positive but did not reach statistical significance (*r* = 0.22, *p* = 0.136). As with the MoCA analysis, this is consistent with range restriction in a clinically homogeneous MCI cohort, where compressed score variability attenuates correlation coefficients. Nevertheless, the positive direction of the correlation for Mini-BESTest, compared to the weak negative association observed for MoCA, may suggest that passively collected movement features bear a more direct relationship with balance and mobility than with cognitive function, though this comparison should be interpreted cautiously given that neither correlation reached statistical significance.

### Feature Importance Analysis

SHAP analysis revealed distinct feature importance profiles for the two clinical assessments. For the MoCA total score, the most influential features were statistical and time-frequency descriptors of the vertical acceleration signal, including absolute sum of changes, mean absolute change, autocorrelation at lag 1, and FFT mean. In contrast, the Mini-BESTest total score was predominantly driven by temporal gait features, with cadence emerging as the single most influential feature by a substantial margin, followed by direction change, stance time, and swing time. This divergence suggests that cognitive and balance functions, while both assessable through passive IMU monitoring, are captured through fundamentally different properties of the movement signal.

#### MoCA

For the MoCA total score, the top contributing features were uniformly related to movement intensity and moment-to-moment fluctuation of the vertical acceleration signal. As shown in Fig 7A, the selected features, such as absolute energy, RMS Vertical Acceleration, absolute sum of changes, and mean absolute change, consistently showed the same directional pattern, where lower values were associated with negative SHAP contributions. Reduced signal energy and fluctuation suggest that the participants with lower MoCA scores might have exhibited reduced movement dynamics. These patterns are consistent with the common behavioral changes reported in the MCI population, such as reduced physical capacity and diminished executive control [23, 76, 77]. More directly, Sakal *et al.* [39] reported that reduced activity intensity was associated with greater risk of poor cognition in free-living accelerometer data, a directional pattern consistent with our finding. Beyond this directional consistency, the most influential features were general movement characteristics rather than gait-specific markers. A waist-mounted IMU continuously captures the whole-body movement across walking and non-walking activities, which together comprise the daily behavioral profile of participants in this therapeutic setting. General intensity and fluctuation features summarize this full profile, and their dominance over gait-specific features in our model suggests that cognitively relevant movement signals in this population were distributed across the broader behavioral spectrum rather than walking pattern alone. This pattern also extends a line of work that reported movement and social interaction features as significant when differentiating higher and lower-functioning MCI cohorts via a camera-based monitoring network in the same therapeutic space [23]. While that work uses a different modality, feature set, and analysis target, our findings add to the broader interpretation that naturalistic behavior in MCI populations within such settings carries information about cognitive function. Future work could explore whether jointly modeling wearable and camera-based features improves subject-level estimation in a naturalistic setting. While the total score analysis highlighted general signal descriptors over gait timing features, the picture at the subdomain level is more mixed. The SHAP analysis (S5 Fig and S6 Fig) showed that the feature importance profiles varied across cognitive subdomains, with temporal gait features such as swing time appearing among the top contributors in several subdomains despite being absent from the total score’s top features. Given the limited sample size for subdomain analysis, however, these patterns should be considered exploratory and require validation in larger cohorts before drawing conclusions about domain-specific associations between gait timing and cognitive function.

#### Mini-BESTest

For the Mini-BESTest total score, cadence was the single most influential feature by a large margin, followed by a combination of temporal gait features such as direction change and stance time, alongside statistical signal descriptors such as mean absolute change and absolute sum of changes. These features directly represent gait rhythm and temporal coordination, and impairments in them reflect deficits in motor control, balance, and executive function that are characteristic of MCI [25, 77]. Maintaining rhythmic gait requires integrated sensorimotor processing and sustained attentional control [27, 76], which explains why these timing features carry signals about both balance and cognitive function. Their prominence aligns with the Mini-BESTest’s emphasis on dynamic postural control, where successful performance depends on efficient weight transfer, anticipatory adjustments, and the ability to sustain stable gait under changing task demands [8]. Beyond this association, our results extend prior gait research in MCI, which has mostly focused on diagnostic group classification (e.g. NC vs MCI vs AD), by showing that the temporal gait features such as cadence and stance time carry sufficient signal to estimate continuous Mini-BESTest scores within MCI. This positions these parameters not only as discriminative markers between cognitive groups but also as quantitative indicators of individual differences in balance function within the MCI population. Notably, this feature importance profile differed substantially from MoCA regression, which placed more weight on general movement intensity and fluctuation. For MCI populations, where cognitive and motor symptoms can progress at different rates and in different combinations, this difference between cognitive and balance feature profiles may be particularly informative for capturing individual-level functional profiles rather than collapsing them into a single global measure. For Mini-BESTest subdomain regressions, the SHAP analysis (S7 Fig and S8 Fig) revealed a separation between subdomains driven primarily by cadence, such as Anticipatory and Sensory Orientation, and subdomains driven by gait-cycle phase timing features, such as swing time and stance time, as observed in both Reactive Postural Control subdomains. However, as with MoCA, these subdomain-level patterns should be considered exploratory given the limited sample size and require validation in larger cohorts.

### Demographic Bias Analysis and Mitigation

Because these models are positioned for potential mobile health (mHealth) use (e.g., passive monitoring, screening augmentation, or triage support), it is important to evaluate whether the model can quantify MoCA and Mini-BESTest equitably across demographic groups. Model shifts depending on demographic groups can translate into unequal downstream decisions, even when overall RMSE is acceptable [28].

#### MoCA

For the sex attribute, bias was present across the MoCA total score and all subdomains before bias mitigation, with ASD values ranging from −0.207 (Attention/Concentration) to 0.292 (Total). After Wasserstein Barycenter mitigation, the total score ASD decreased substantially from 0.292 to −0.010 and SP AUC from 0.143 to 0.033, while RMSE remained stable (3.294 to 3.291). SP AUC was consistently reduced across all subdomains, with post-mitigation values ranging from 0.074 (Memory) to 0.100 (Executive). Notably, Language (ASD: −0.122 to −0.047) and Visuospatial (ASD: 0.168 to −0.030) reached near-zero ASD after bias mitigation, suggesting approximately equitable performance for males and females in these domains. These results indicate that the Wasserstein Barycenter method can effectively reduce sex-related disparities when demographic groups are balanced (22 males and 22 females).

For the race attribute, pre-mitigation ASD values were already close to zero for the total score (−0.003) and several subdomains including Memory (0.003), Language (−0.019), and Orientation (−0.004), suggesting relatively low initial bias. However, bias mitigation produced adverse effects across most targets, with the total score ASD increasing from −0.003 to 0.172 and similar overcorrections observed for Attention/Concentration (0.037 to 0.274), Executive (−0.149 to 0.209), and Memory (0.003 to 0.118). Nevertheless, SP AUC for race showed modest reductions for most subdomains (e.g., Visuospatial: 0.268 to 0.155) except for Memory and Attention/Concentration scores. This suggests that the predicted score distributions between racial groups became more similar overall, despite the mean scores diverging as reflected by increased ASD. The increased biases even after bias mitigation could be attributed to a severe imbalance between White and Black/African American participants, where we had 6.33 times larger number of White participants. Previous studies have demonstrated that underrepresentation and unbalanced subgroup strata introduce bias and uncertainty in ML models [28], highlighting the importance of balanced demographic ratios for reliable bias mitigation, as supported by our bias mitigation analysis of the sex attribute, where equal group sizes enabled effective correction. Future work should collect balanced samples from various racial groups to further study biases in wearable-based ML models for MCI populations and the feasibility of mitigating these biases.

#### Mini-BESTest

Unlike the MoCA results where sex-related bias varied in direction across subdomains, the Mini-BESTest models exhibited a consistent pattern of systematically underestimating scores for the female group, with negative ASD values across the total score (−0.414) and all subdomains before bias mitigation. Wasserstein Barycenter mitigation effectively corrected this pattern, reducing the total score ASD to −0.009 and SP AUC from 0.121 to 0.034, with RMSE preserved (3.529 to 3.496). Across subdomains, all post-mitigation SP AUC values fell below 0.042, and several targets including Reactive Postural Control Right (−0.144 to −0.006), Reactive Postural Control Left (−0.072 to −0.005), and Anticipatory Right (−0.123 to −0.005) reached near-zero ASD. This effective mitigation is supported by the nearly balanced representation of males and females in our participants with Mini-BESTest scores (19 males and 22 females). For the race attribute, our bias mitigation improved fairness for Sensory Orientation (ASD: −0.244 to 0.029) and Reactive Postural Control Right (ASD: 0.126 to 0.014), but overcorrected for Anticipatory Right (ASD: −0.166 to 0.154) and Anticipatory Left (ASD: − 0.049 to 0.173). The total score showed the most pronounced overcorrection, with ASD reversing from −0.439 to 0.537 despite SP AUC improving from 0.175 to 0.117. Like MoCA, these inconsistencies reflect the severe demographic imbalance of 35 White participants and 6 Black / African American participants. This again highlights the need for more demographically diverse cohorts to draw reliable conclusions on race-related bias mitigation in future studies.

### Potential Clinical Application

From a care delivery perspective, these models motivate the continued investigation of wearable-based remote passive monitoring of movement characteristics during everyday activities, as this addresses a fundamental limitation of episodic clinic-based assessments by providing an ecologically valid view of function between encounters [78]. This may be particularly relevant in settings where in-person testing is burdensome or infrequent. The divergence in feature importance profiles between the two assessments suggests potential value for triage and personalization. For instance, individuals whose gait exhibits changes in gait cadence and phase timing may benefit from targeted balance-focused evaluation and rehabilitation [79], whereas individuals whose movement patterns show reduced overall movement intensity may warrant closer attention to cognitive screening [80]. Future clinical translation should therefore emphasize longitudinal within-person change and prospective validation of whether movement-derived and gait-derived features predict clinically meaningful endpoints such as functional decline, falls, or response to intervention.

## Limitation

CEP includes multiple functional spaces (e.g., gym, multipurpose rooms, kitchen) designed to facilitate the natural interactions and movements of free-living environments. Our study design also shows that wearable sensing can quantify movements in MCI within such settings. However, it remains far from a home environment, and participants may not show the behaviors present in their daily living. For example, our dataset does not include walking on uneven terrain or walking up and down the stairs. Our future work will evaluate the ecological validity of the trained model by sending wearables and AI models to quantify behaviors in MCI in daily living [39, 40] by continually adapting the AI model to personalize it [81, 82] for each participant’s home and daily living settings. We expect this future study to reveal day-to-day fluctuations in cognitive and balance functions that have significant implications in the quality of life for older adults with MCI. Beyond day-to-day fluctuations, the current dataset can support analysis of within-programs during 6-month periods of the 6-month CEP intervention. The future study will deploy wearable sensors to participants during the post-intervention period after they exit CEP to assess changes in gait features associated with cognitive impairment and balance functions in daily living following lifestyle interventions. In addition, our FFT-based features summarize the overall magnitude and spread of spectral energy rather than decomposing the gait-relevant frequency range into physiologically meaningful sub-bands (e.g., stride frequency, step frequency, or harmonics). Future work will extend the feature set with sub-band spectral descriptors to more precisely characterize frequency content associated with gait-related movement patterns in MCI. Our study showed that the wearable AI models naively trained on our dataset were biased toward sex and race attributes, despite the balanced number of female and male participants. The diverse populations with various profiles of cognitive decline will provide an excellent benchmark for studying the equitable wearable AI for cognitive impairments to maximize the ecological validity in marginalized communities with limited access to dementia care [83]. Our modest sample size (N=44) and limited racial diversity, with 85% of participants identifying as White, may not fully capture the heterogeneity of MCI and limit the strength of conclusions about model fairness across demographic groups. Future work should evaluate these models in larger and more racially and ethnically diverse cohorts spanning normal cognition, MCI, and early ADRD to assess their generalizability and equitable performance across the continuum of early cognitive decline.

## Conclusion

With the growing prevalence of dementia and disparities in access to dementia care [1, 2], there is an urgent need to develop scalable monitoring systems for older adults with MCI that can inform personalized intervention. Wearable sensing systems offer opportunities for passive, unobtrusive, and continuous monitoring of behaviors in daily lives, but previous studies were largely limited to controlled laboratory settings. This study addressed this gap and demonstrated the feasibility of an equitable AI-driven wearable sensing system for quantifying cognitive and balance functions in older adults with MCI during natural movements and social interactions, as assessed by the MoCA and Mini-BESTest scores. Our models achieved RMSE of 3.677 and 3.672 for regressing MoCA and Mini-BESTest total scores, respectively. The RMSE for MoCA regression was within the MDC of 5.1, and the RMSE for Mini-BESTest regression, while slightly above the MDC of 3.4, remained below the MCID of 3.8. Furthermore, our model could reliably quantify the subdomains of cognitive and balance functions, demonstrating its potential for personalized interventions. Our feature importance analysis revealed that the two assessments relied on distinct properties of the movement signal, with general movement intensity features most informative for MoCA and temporal gait features led by cadence most informative for Mini-BESTest. This suggests that both broader movement characteristics and biomechanical gait markers may serve as complementary signals for ecologically valid and proxy measurement of cognitive and balance function in everyday settings, providing wearables as accessible and scalable monitoring tools. Our bias analysis showed that wearable ML models are biased for both sex and race attributes, and that Wasserstein Barycenter bias mitigation can reduce biases on sex attribute while maintaining model performance. This analysis also showed that the imbalanced ratio of race representation in our dataset makes it challenging to mitigate data bias induced in the model. Taken together, this work sets an important milestone for future research on wearable sensing for longitudinal monitoring of cognitive and balance functions in older adults with cognitive impairment, with diverse clinical and demographic representation, in naturalistic environments.

## Data Availability

Requests for data access may be directed to Dr. Hyeokhyen Kwon or the Emory Department of Biomedical Informatics.

## Acknowledgments

Hyeokhyen Kwon, Rachel Hershenberg, Bolaji Omofojoye, and Gari Clifford are partially funded by the National Institute on Deafness and Other Communication Disorders (grant # 1R21DC021029-01A1). The Cognitive Empowerment Program is supported by a generous investment from the James M. Cox Foundation and Cox Enterprises, Inc. We thank Dr. Allan I. Levey (Department of Neurology, Emory University) for providing thoughtful feedback and valuable contributions to this study.

## Supporting information

**S1 Table.** Extracted features from accelerometer and gyroscope data. This table defines the accelerometer-and gyroscope-derived features used for model training.

| Feature name | Definition |
| --- | --- |
| abs energy | Calculates the absolute energy of the filtered vertical acceleration signal, which is the sum of its squared values. |
| absolute maximum | Calculates the maximum absolute value observed in the filtered vertical acceleration signal. |
| absolute sum of changes | Calculates the sum of the absolute values of consecutive differences between data points in the filtered vertical acceleration signal. |
| autocorrelation lag1 | Calculates the Pearson correlation coefficient between the filtered vertical acceleration signal and itself shifted by one time step. |
| binned entropy | Calculates the Shannon entropy based on the probability distribution from a 10-bin histogram of the filtered vertical acceleration signal's values. |
| cadence | Calculates the number of steps per minute, derived from the count of peaks found in the filtered vertical acceleration signal. |
| change quantiles | Calculates the interquartile range (IQR) of the filtered vertical acceleration signal, defined as the 75th percentile minus the 25th percentile. |
| direction change | Calculates the mean of the absolute consecutive differences in the gyroscope signal magnitude, indicating the average rate of change in angular velocity. |
| ECDF 0 | The 0th percentile, or minimum value, of the filtered vertical acceleration signal. |
| ECDF 5 | The 5th percentile of the filtered vertical acceleration signal. |
| ECDF 25 | The 25th percentile, or first quartile, of the filtered vertical acceleration signal. |
| ECDF 50 | The 50th percentile, or median, of the filtered vertical acceleration signal. |
| ECDF 75 | The 75th percentile, or third quartile, of the filtered vertical acceleration signal. |
| ECDF 95 | The 95th percentile of the filtered vertical acceleration signal. |
| ECDF 100 | The 100th percentile, or maximum value, of the filtered vertical acceleration signal. |
| ECDF mean | The arithmetic mean of the filtered vertical acceleration signal. |
| ECDF std | The standard deviation of the filtered vertical acceleration signal. |
| entropy orientation | Calculates the Shannon entropy from a histogram of the gyroscope signal magnitude, reflecting the predictability of orientation changes. |
| entropy vertical acceleration | Calculates the Shannon entropy from a histogram of the filtered vertical acceleration signal, reflecting the predictability of vertical movement. |
| fft mean | Calculates the arithmetic mean of the absolute values of the Fast Fourier Transform (FFT) coefficients of the filtered vertical acceleration signal. |
| fft var | Calculates the variance of the absolute values of the FFT coefficients of the filtered vertical acceleration signal. |
| fourier entropy | Calculates the Shannon entropy from the first 10 components of the power spectral density (PSD) obtained using Welch's method. |
| gait velocity | Calculates walking speed, derived by dividing stride length by stride duration, where both are computed from the same initial contact events identified by the Zijlstra and Hof (2003) method. |
| kurtosis | Calculates the kurtosis, a measure of the tailedness or sharpness of the peak, of the filtered vertical acceleration signal distribution. |
| linear trend intercept | Calculates the intercept, or value at time 0, of a linear regression line fitted to the filtered vertical acceleration signal values over time. |
| linear trend slope | Calculates the slope of a linear regression line fitted to the filtered vertical acceleration signal values, indicating the overall trend. |
| longest strike above mean | Calculates the length of the longest consecutive sequence of data points that are above the arithmetic mean of the filtered vertical acceleration signal. |
| mean abs change | Calculates the average of the absolute values of consecutive differences between data points in the filtered vertical acceleration signal. |
| mean second derivative central | Calculates the mean of the second-order differences of the filtered vertical acceleration signal, approximating the average central second derivative. |
| number peaks | Calculates the total count of peaks in the filtered vertical acceleration signal, where peaks are required to be separated by at least 11 samples, corresponding to approximately 0.5 seconds at 23 Hz. |
| RMS jerk | Calculates the root mean square (RMS) of jerk, which is the time derivative of the signal vector magnitude (SVM) of the filtered accelerometer data. |
| RMS vertical acceleration | Calculates the root mean square (RMS) of the filtered vertical acceleration signal. |
| skewness | Calculates the skewness, a measure of asymmetry, of the filtered vertical acceleration signal probability distribution. |
| stance time | Estimates the time the foot is on the ground, calculated as 60% of the average stride duration. |
| gait symmetry index | Quantifies step time asymmetry as the absolute difference between mean left and mean right step durations, divided by their average and expressed as a percentage. |
| stride duration | Calculates the average time for a full gait cycle, or two steps, derived from the time between consecutive step peaks. |
| stride length | Calculates stride length using the inverted pendulum model of Zijlstra and Hof (2003), where initial contacts are detected from anterior-posterior acceleration zero-crossings and vertical displacement is obtained by double integration of vertical acceleration with high-pass drift correction. |
| stride time CV | Calculates the coefficient of variation, defined as standard deviation divided by mean, of the stride durations and expresses it as a percentage. |
| stride variability | Calculates the coefficient of variation, defined as standard deviation divided by mean, of the stride durations and expresses it as a decimal proportion. |
| swing time | Estimates the time the foot is in the air, calculated as 40% of the average stride duration. |

---

**S1 Fig.**
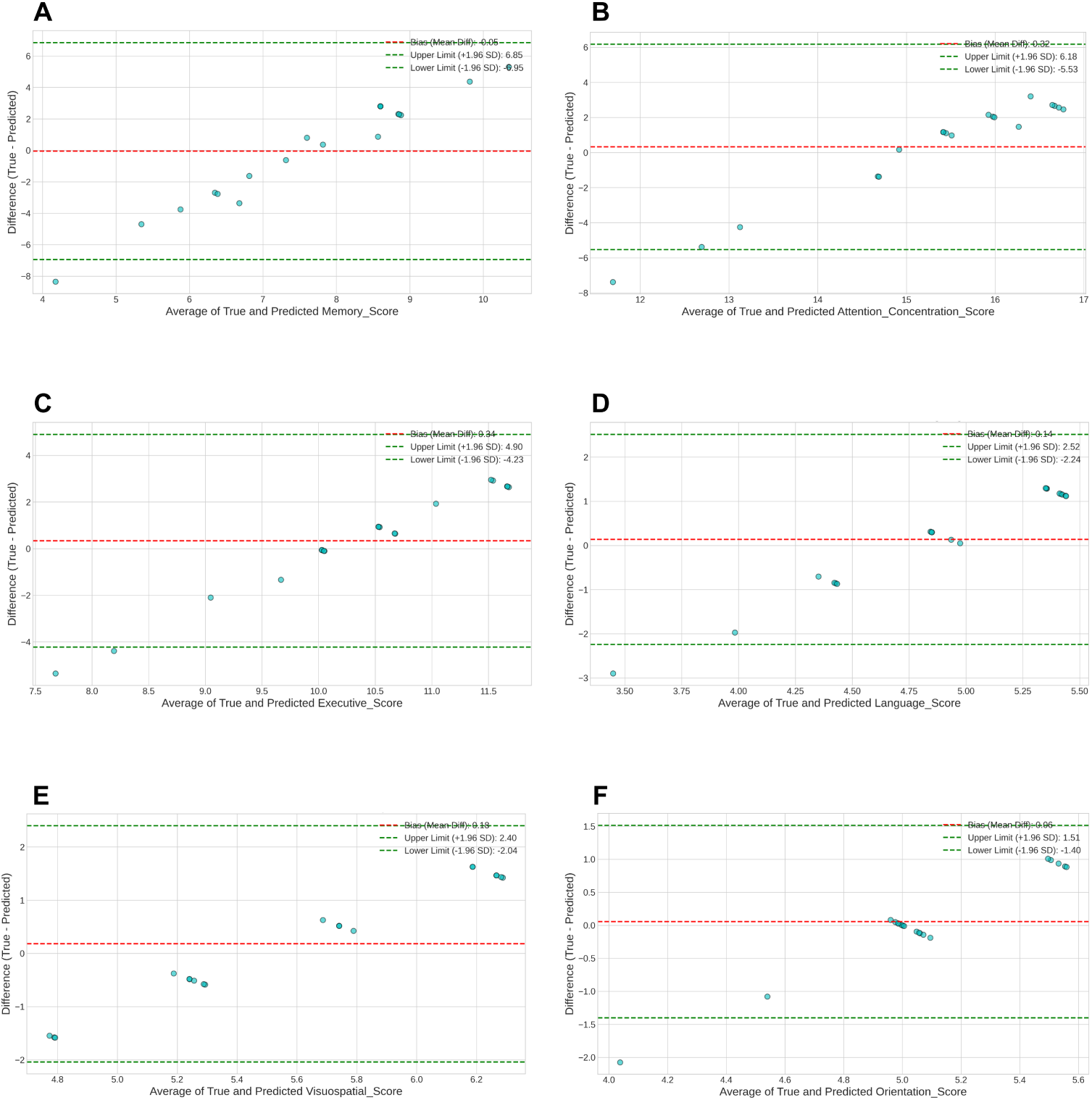
Bland–Altman plots for MoCA subdomain score regressions. A: Memory. B: Attention/concentration. C: Executive. D: Language. E: Visuospatial. F: Orientation. The dashed lines indicate mean bias and 95% limits of agreement.

**S2 Fig.**
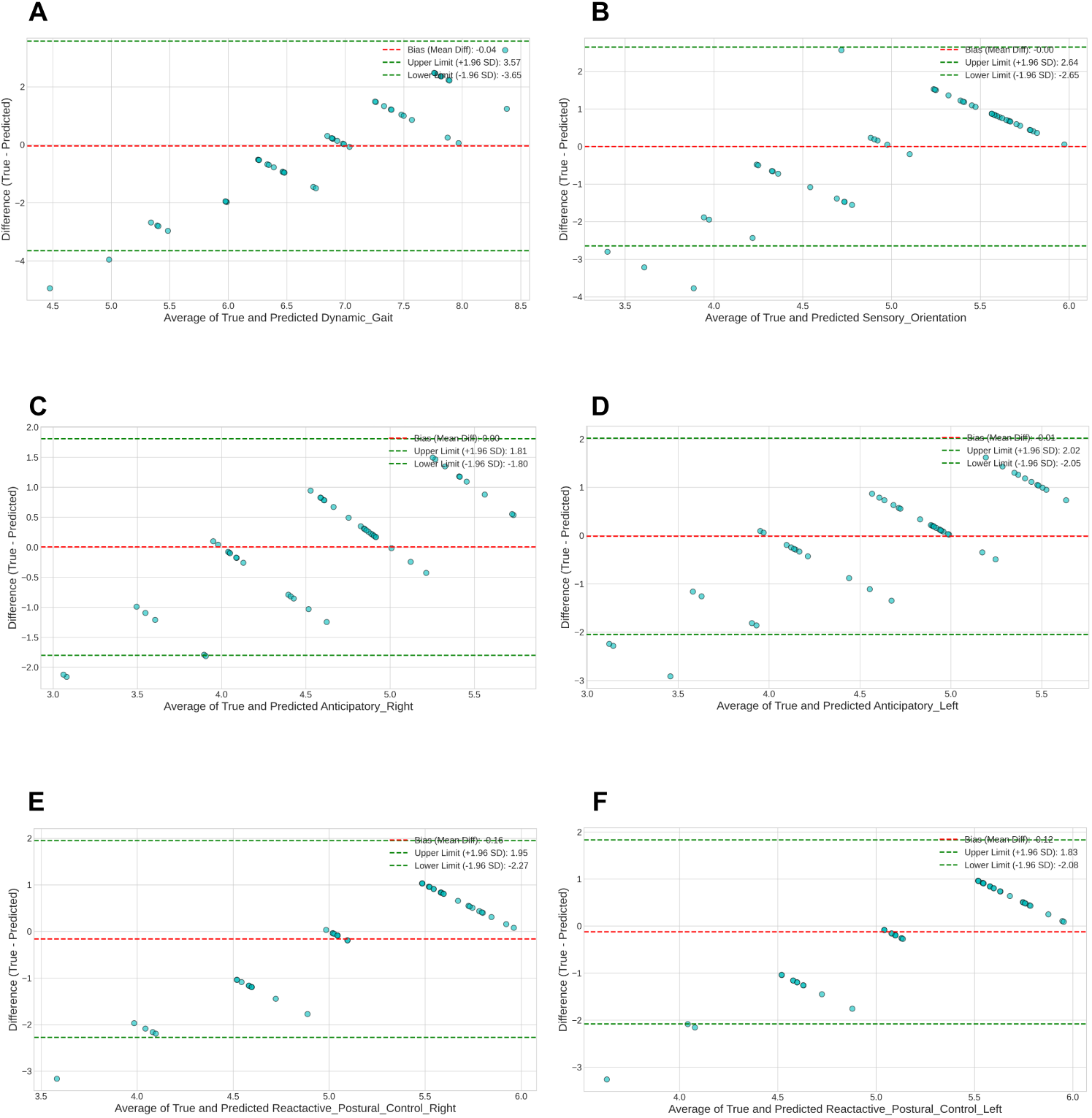
Bland–Altman plots for Mini-BESTest subdomain score regressions. A: Dynamic gait. B: Sensory orientation. C: Reactive postural control right. D: Reactive postural control left. E: Anticipatory right. F: Anticipatory left. The dashed lines indicate mean bias and 95% limits of agreement.

**S3 Fig.**
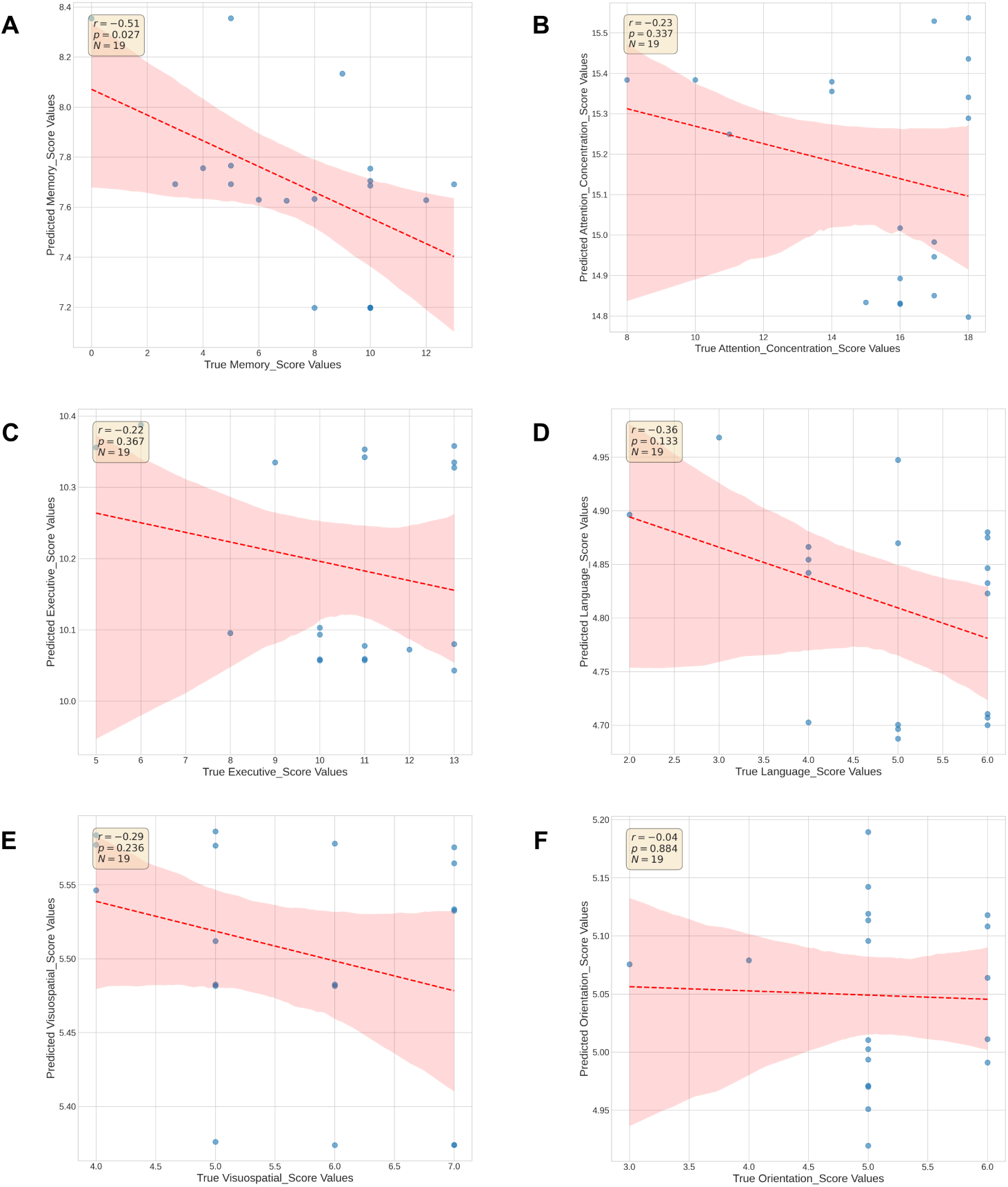
Pearson correlation plots for MoCA subdomain score regressions. A: Memory. B: Attention/concentration. C: Executive. D: Language. E: Visuospatial. F: Orientation. The regression line and 95% confidence band are shown for each subdomain.

**S4 Fig.**
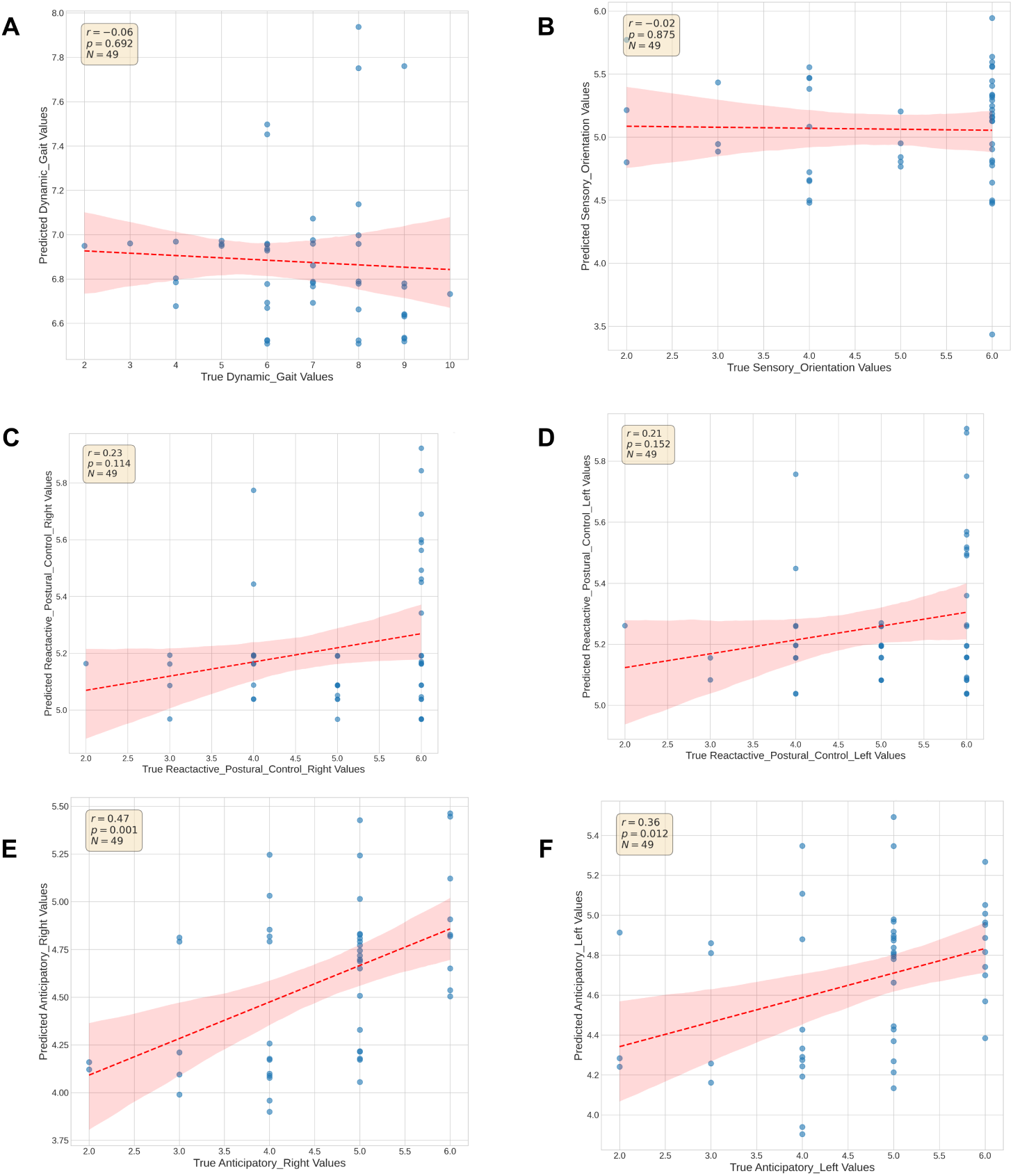
Pearson correlation plots for Mini-BESTest subdomain score regressions. A: Dynamic gait. B: Sensory orientation. C: Reactive postural control right. D: Reactive postural control left. E: Anticipatory right. F: Anticipatory left. The regression line and 95% confidence band are shown for each subdomain.

**S5 Fig.**
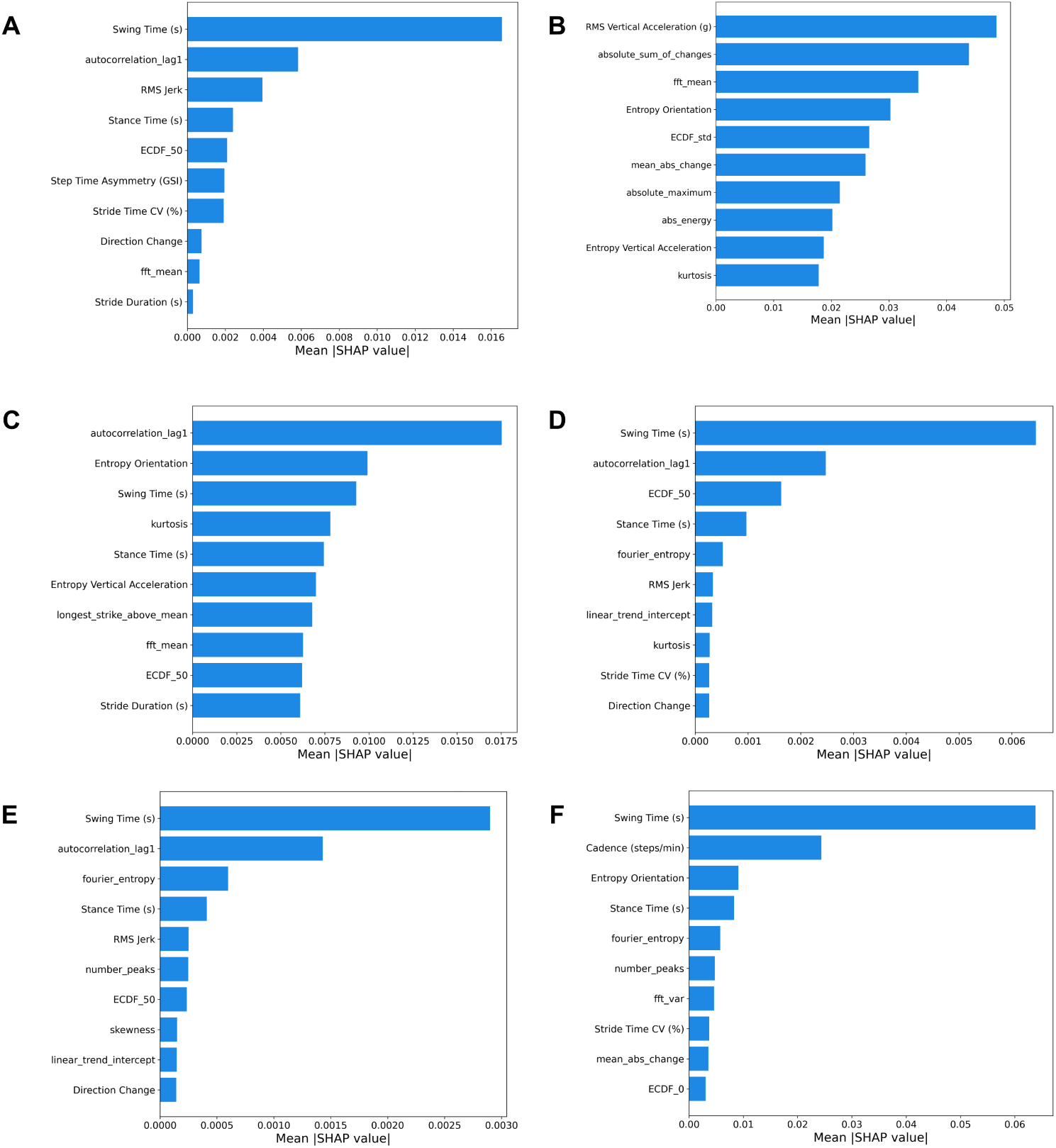
SHAP feature importance for MoCA subdomain score regressions. A: Memory. B: Attention/concentration. C: Executive. D: Language. E: Visuospatial. F: Orientation. The plots show mean absolute SHAP values for the random forest models trained to predict each MoCA subdomain score.

**S6 Fig.**
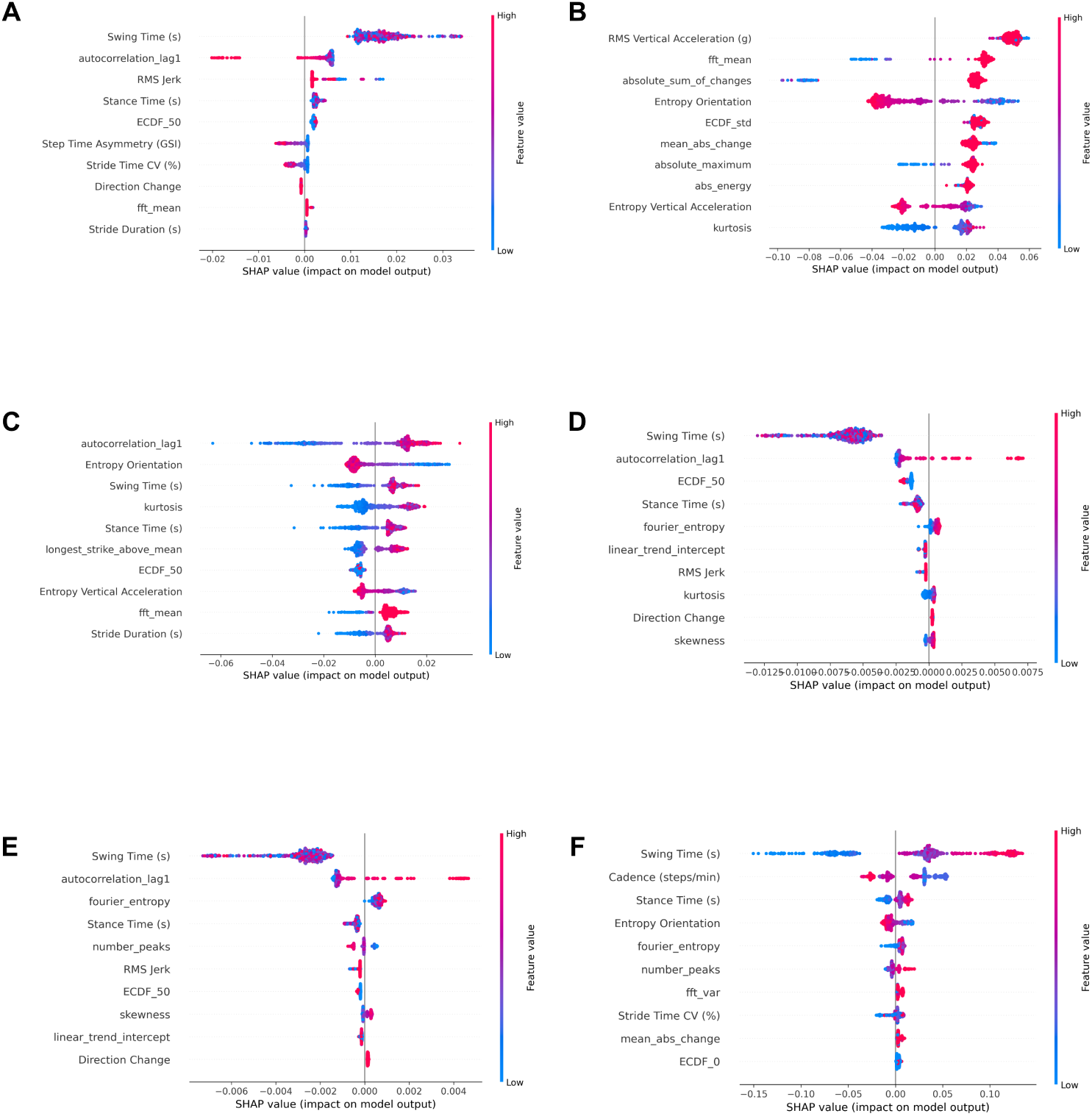
SHAP beeswarm plots for MoCA subdomain score regressions. A: Memory. B: Attention/concentration. C: Executive. D: Language. E: Visuospatial. F: Orientation. The plots show SHAP value distributions for the random forest models trained to predict each MoCA subdomain score.

**S7 Fig.**
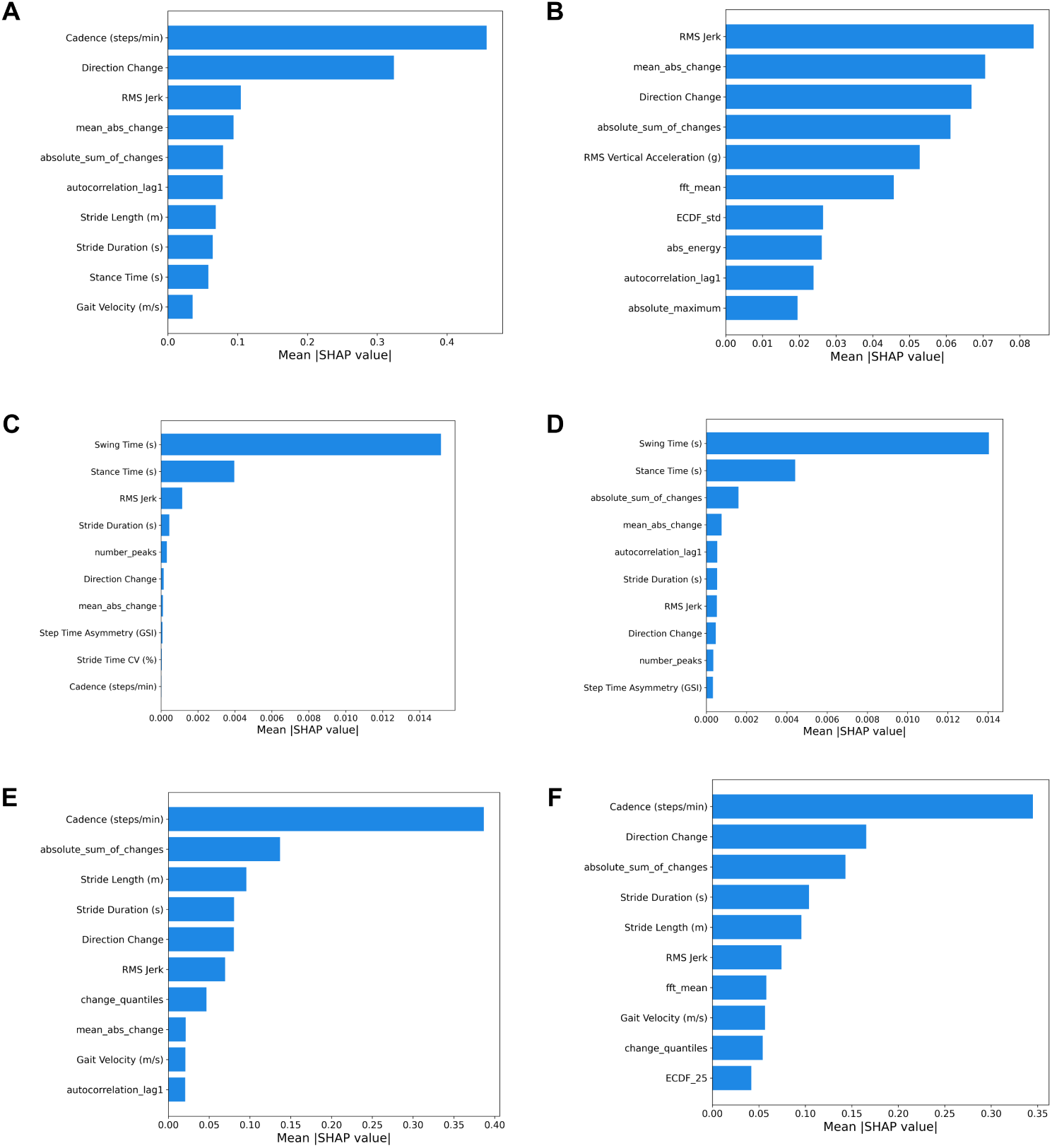
SHAP feature importance for Mini-BESTest subdomain score regressions. A: Dynamic gait. B: Sensory orientation. C: Reactive postural control right. D: Reactive postural control left. E: Anticipatory right. F: Anticipatory left. The plots show mean absolute SHAP values for the best-performing model trained to predict each Mini-BESTest subdomain score.

**S8 Fig.**
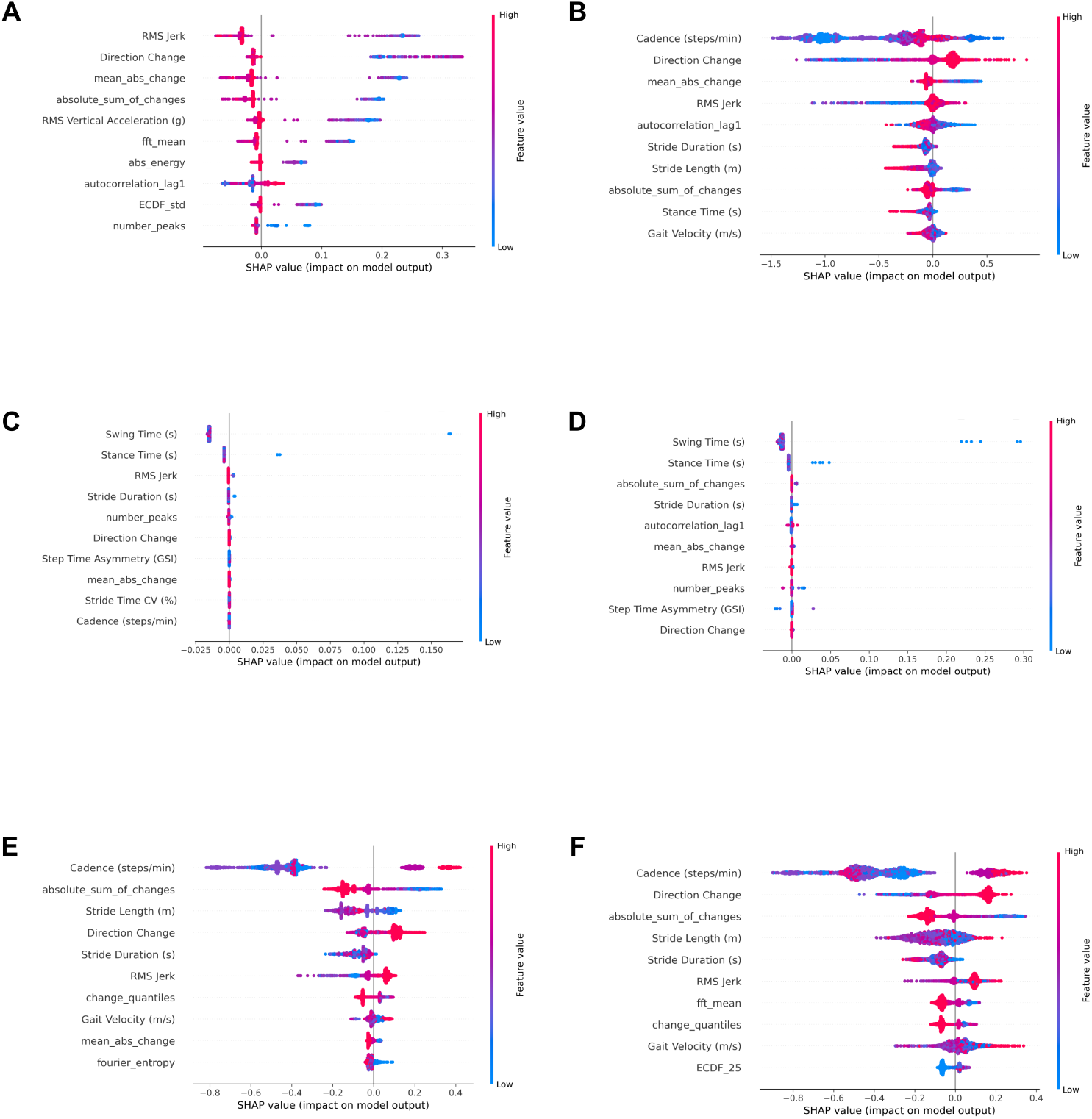
SHAP beeswarm plots for Mini-BESTest subdomain score regressions. A: Dynamic gait. B: Sensory orientation. C: Reactive postural control right. D: Reactive postural control left. E: Anticipatory right. F: Anticipatory left. The plots show SHAP value distributions for the best-performing model trained to predict each Mini-BESTest subdomain score.

## Footnotes

1 https://yostlabs.com/product/3-space-data-logger/

## References

1. Organization WH. Dementia Fact Sheet. WHO. 2021. Available from: https://www.who.int/news-room/fact-sheets/detail/dementia.

2. (ADI) ADI. Dementia statistics: Facts and Figures. WHO. 2021. Available from: https://www.alzint.org/about/dementia-facts-figures/dementia-statistics/#:~:text=,dementia.

3. Negash S, Petersen RC. Mild cognitive impairment: an overview. CNS Spectrums. 2008;13(1):45–53. doi:10.1017/S1092852900016151.

4. Seifallahi M, Galvin JE, Ghoraani B. Detection of mild cognitive impairment using various types of gait tests and machine learning. Frontiers in Neurology. 2024;15. doi:10.3389/fneur.2024.1354092.

5. Nasreddine ZS, Phillips NA, Bédirian V, Charbonneau S, Whitehead V, Collin I, et al. The Montreal Cognitive Assessment, MoCA: A Brief Screening Tool For Mild Cognitive Impairment. Journal of the American Geriatrics Society. 2005;53(4):695–9. Available from: https://agsjournals.onlinelibrary.wiley.com/doi/abs/10.1111/j.1532-5415.2005.53221.x.arXiv: https://agsjournals.onlinelibrary.wiley.com/doi/pdf/10.1111/j.1532-5415.2005.53221.x. 10.1111/j.1532-5415.2005.53221.x.

6. Bahureksa L, Najafi B, Saleh A, Sabbagh M, Coon D, Mohler MJ, et al. The Impact of Mild Cognitive Impairment on Gait and Balance: A Systematic Review and Meta-Analysis of Studies Using Instrumented Assessment. Gerontologia. 2016 May;63(1):67–83. arXiv:https://karger.com/ger/article-pdf/63/1/67/2838340/000445831.pdf. doi:10.1159/000445831.

7. Bovonsunthonchai S, Vachalathiti R, Hiengkaew V, Bryant MS, Richards J, Senanarong V. Quantitative Gait Analysis in Mild Cognitive Impairment, Dementia, and Cognitively Intact Individuals: A Cross-Sectional Case–Control Study. BMC Geriatrics. 2022 Sep;22(1):767. doi:10.1186/s12877-022-03405-9.

8. Franchignoni F, Horak F, Godi M, Nardone A, Giordano A. Using psychometric techniques to improve the Balance Evaluation Systems Test: the mini-BESTest. Journal of Rehabilitation Medicine. 2010 Mar;42(4):323–331. Available from: https://medicaljournalssweden.se/jrm/article/view/17823. doi:10.2340/16501977-0537.

9. Langa KM, Levine DA. The diagnosis and management of mild cognitive impairment: a clinical review. JAMA. 2014;312(23):2551–61. doi:10.1001/jama.2014.13806.

10. Xu Q, Kim Y, Chung K, Schulz P, Gottlieb A. Prediction of Mild Cognitive Impairment Status: Pilot Study of Machine Learning Models Based on Longitudinal Data From Fitness Trackers. JMIR Form Res. 2024. doi:10.2196/55575.

11. Liu JL, Baker L, Chen AYA, Wang JJ. Geographic Variation in Shortfalls of Dementia Specialists in the United States. Health Affairs Scholar. 2024 Jul;2(7):qxae088. doi:10.1093/haschl/qxae088.

12. Amiri S, Keffeler JI, Crain DR, Denney JT, Buchwald D. Racial, ethnic, and rural disparities in distance to physicians among decedents with Alzheimer’s disease and related dementias in Washington State. Alzheimer’s & Dementia. 2024;20(5):3671–8. Available from: https://alz-journals.onlinelibrary.wiley.com/doi/abs/10.1002/alz.13756. arXiv:https://alz-journals.onlinelibrary.wiley.com/doi/pdf/10.1002/alz.13756. 10.1002/alz.13756.

13. Singh S, Strong R, Xu I, Fonseca LM, Hawks Z, Grinspoon E, et al. Ecological Momentary Assessment of Cognition in Clinical and Community Samples: Reliability and Validity Study. Journal of Medical Internet Research. 2023 Jun;25(1):e45028. doi:10.2196/45028.

14. Leach JM, Mancini M, Kaye JA, Hayes TL, Horak FB. Day-to-Day Variability of Postural Sway and Its Association With Cognitive Function in Older Adults: A Pilot Study. Frontiers in Aging Neuroscience. 2018 May;10:126. doi:10.3389/fnagi.2018.00126.

15. Mohr DC, Zhang M, Schueller SM. Personal Sensing: Understanding Mental Health Using Ubiquitous Sensors and Machine Learning [Journal Article]. Annual Review of Clinical Psychology. 2017;13(Volume 13, 2017):23-47. doi:10.1146/annurev-clinpsy-032816-044949.

16. Verghese J, Robbins M, Holtzer Rea. Gait dysfunction in mild cognitive impairment syndromes. J Am Geriatr Soc. 2008;56(7):1244–51. doi:10.1111/j.1532-5415.2008.01758.x.

17. Boettcher LN, Hssayeni M, Rosenfeld A, Tolea MI, Galvin JE, Ghoraani B. Dual-Task Gait Assessment and Machine Learning for Early-detection of Cognitive Decline. In: 2020 42nd Annual International Conference of the IEEE Engineering in Medicine & Biology Society (EMBC); 2020. p. 3204–7. doi:10.1109/EMBC44109.2020.9175955.

18. Xie H, Wang Y, Tao S, Huang S, Zhang C, Lv Z. Wearable Sensor-Based Daily Life Walking Assessment of Gait for Distinguishing Individuals With Amnestic Mild Cognitive Impairment. Frontiers in Aging Neuroscience. 2019 Oct;11. doi:10.3389/fnagi.2019.00285.

19. Zhao H, Cao J, Xie J, Liao WH, Lei Y, Cao H, et al. Wearable Sensors and Features for Diagnosis of Neurodegenerative Diseases: A Systematic Review. DIGITAL HEALTH. 2023 Jan;9:20552076231173569. doi:10.1177/20552076231173569.

20. Jamshed M, Shahzad A, Riaz Fea. Exploring inertial sensor-based balance biomarkers for early detection of mild cognitive impairment. Sci Rep. 2024;14(9829). doi:10.1038/s41598-024-59928-1.

21. Warmerdam E, Hausdorff JM, Atrsaei A, Zhou Y, Mirelman A, Aminian K, et al. Long-term unsupervised mobility assessment in movement disorders. The Lancet Neurology. 2020;19(5):462–70. Available from: https://www.sciencedirect.com/science/article/pii/S1474442219303977. doi: 10.1016/S1474-4422(19)30397-7.

22. Jeon Y, Kang J, Kim BS, Lee KH, Song J, Gwak J. Early Alzheimer’s disease diagnosis using wearable sensors and multilevel gait assessment: a machine learning ensemble approach. IEEE Sensors Journal. 2023;23(9):10041–53. doi:10.1109/JSEN.2023.3259034.

23. Hegde C, Kiarashi Y, Levey AI, Rodriguez AD, Kwon H, Clifford GD. Feasibility of assessing cognitive impairment via distributed camera network and privacy-preserving edge computing. Alzheimer’s & Dementia: Diagnosis, Assessment & Disease Monitoring. 2025;17(1):e70085. Available from: https://alz-journals.onlinelibrary.wiley.com/doi/abs/10.1002/dad2.70085. arXiv:https://alz-journals.onlinelibrary.wiley.com/doi/pdf/10.1002/dad2.70085. 10.1002/dad2.70085.

24. Ghoraani B, Boettcher LN, Hssayeni MD, Rosenfeld A, Tolea MI, Galvin JE. Detection of mild cognitive impairment and Alzheimer’s disease using dual-task gait assessments and machine learning. Biomedical Signal Processing and Control. 2021;64:102249. Available from: https://www.sciencedirect.com/science/article/pii/S1746809420303773. doi: 10.1016/j.bspc.2020.102249.

25. Lindh-Rengifo M, Jonasson SB, Ullén S, Stomrud E, Palmqvist S, Mattsson-Carlgren N, et al. Components of Gait in People with and without Mild Cognitive Impairment. Gait & Posture. 2022;93:83–9. doi:10.1016/j.gaitpost.2022.01.012.

26. Masse FAA, Ansai JH, Gerassi RC, Tsen C, de Castro Cezar NO, de Andrade LP. Six-Month Change in Gait Speed to Discriminate between Those with and without Falls History in Older People with Mild Cognitive Impairment and Mild Alzheimer Disease. Geriatric Nursing. 2022;48:274–9. doi:10.1016/j.gerinurse.2022.10.002.

27. Saini M, Subramanian MS, Rao AR, Thakral M, Singh V, Chakrawarty A, et al. Gait Parameters Change Can Be an Early Marker of Cognitive Impairment. Neurology India. 2024 May;72(3):603–9. doi:10.4103/ni.ni_148_22.

28. Yuan C, Linn KA, Hubbard RA. Algorithmic Fairness of Machine Learning Models for Alzheimer Disease Progression. JAMA Network Open. 2023 11;6(11):e2342203-3. Available from: doi:10.1001/jamanetworkopen.2023.42203.

29. Yfantidou S, Constantinides M, Spathis D, Vakali A, Quercia D, Kawsar F. (Un)Fair Devices: Moving beyond AI Accuracy in Personal Sensing. ACM Journal on Responsible Computing. 2026 Jun;3(2):1–24. arXiv:2303.15585. doi:10.1145/3796223.

30. Zhong Y, Huang S, Zou M, Chen Y, Shen P, He Y, et al. Gait Analysis in Older Adults with Mild Cognitive Impairment: A Bibliometric Analysis of Global Trends, Hotspots, and Emerging Frontiers. Frontiers in Aging. 2025;Volume 6 - 2025. doi:10.3389/fragi.2025.1592464.

31. Lin J, Xu T, Yang X, Yang Q, Zhu Y, Wan M, et al. A detection model of cognitive impairment via the integrated gait and eye movement analysis from a large Chinese community cohort. Alzheimer’s & Dementia. 2024;20(2):1089–101. Available from: https://alz-journals.onlinelibrary.wiley.com/doi/abs/10.1002/alz.13517. arXiv:https://alz-journals.onlinelibrary.wiley.com/doi/pdf/10.1002/alz.13517. 10.1002/alz.13517.

32. Knapstad MK, Naterstad I, Bogen B. The Association between Cognitive Impairment, Gait Speed, and Walk Ratio. Frontiers in Aging Neuroscience. 2023;Volume 15 - 2023. doi:10.3389/fnagi.2023.1092990.

33. Tolea MI, Rosenfeld A, Roy SV, Besser LM, O’Shea DM, Galvin JE. Gait, Balance, and Physical Performance as Markers of Early Alzheimer’s Disease and Related Dementia Risk. Journal of Alzheimer’s Disease. 2025 Nov;108(1 suppl):S185-99. doi:10.1177/13872877241313144.

34. Longhurst J, Phan J, Chen E, Jackson S, Landers MR. Physical Therapy for Gait, Balance, and Cognition in Individuals with Cognitive Impairment: A Retrospective Analysis. Rehabilitation Research and Practice. 2020;2020(1):8861004. Available from: https://onlinelibrary.wiley.com/doi/abs/10.1155/2020/8861004. arXiv:https://onlinelibrary.wiley.com/doi/pdf/10.1155/2020/8861004. 10.1155/2020/8861004.

35. Cejudo A, Arrojo M, Martín C, Almeida A. AI and Wearables for Early Detection of Cognitive Impairment and Dementia: Systematic Review. Journal of Medical Internet Research. 2026 Feb;28(1):e86262. doi:10.2196/86262.

36. Pau M, Mulas I, Putzu V, Asoni G, Viale D, Mameli I, et al. Smoothness of Gait in Healthy and Cognitively Impaired Individuals: A Study on Italian Elderly Using Wearable Inertial Sensor. Sensors. 2020 Jan;20(12):3577. doi:10.3390/s20123577.

37. Jeon Y, Kang J, Kim BC, Ho Lee K, Song JI, Gwak J. Smart Insole-Based Classification of Alzheimer’s Disease Using Few-Shot Learning Facilitated by Multi-Scale Metric Learning. IEEE Transactions on Consumer Electronics. 2024 May;70(2):4699–708. doi:10.1109/TCE.2024.3386714.

38. Robles Cruz D, Puebla Quiñones S, Lira Belmar A, Quintana Figueroa D, Reyes Hidalgo M, Taramasco Toro C. Fall Risk Classification Using Trunk Movement Patterns from Inertial Measurement Units and Mini-BESTest in Community-Dwelling Older Adults: A Deep Learning Approach. Applied Sciences. 2024 Jan;14(20):9170. doi:10.3390/app14209170.

39. Sakal C, Li T, Li J, Li X. Predicting Poor Performance on Cognitive Tests among Older Adults Using Wearable Device Data and Machine Learning: A Feasibility Study. npj Aging. 2024 Nov;10(1):56. doi:10.1038/s41514-024-00177-x.

40. Chan LLY, Espinoza Cerda MT, Brodie MA, Lord SR, Taylor ME. Daily-Life Walking Speed, Running Duration and Bedtime from Wrist-Worn Sensors Predict Incident Dementia: A Watch Walk – UK Biobank Study. International Psychogeriatrics. 2025 Jun;37(3):100031. doi:10.1016/j.inpsyc.2024.100031.

41. Reinertsen E, Osipov M, Liu C, Kane JM, Petrides G, Clifford GD. Continuous Assessment of Schizophrenia Using Heart Rate and Accelerometer Data. Physiological Measurement. 2017 Jun;38(7):1456. doi:10.1088/1361-6579/aa724d.

42. Suresha PB, O’Leary H, Tarquinio DC, Hehn JV, Clifford GD. Rett Syndrome Severity Estimation with the BioStamp nPoint Using Interactions between Heart Rate Variability and Body Movement. PLOS ONE. 2023 Mar;18(3):e0266351. doi:10.1371/journal.pone.0266351.

43. Rad AB, Villavicencio T, Kiarashi Y, Anderson C, Foster J, Kwon H, et al. From Motion to Emotion: Exploring Challenging Behaviors in Autism Spectrum Disorder through Analysis of Wearable Physiology and Movement. Physiological Measurement. 2025 Jan;46(1):015004. doi:10.1088/1361-6579/ada51b.

44. Rodriguez AD, DuBose JR, Rozga A, Zimring CM, Mynatt ED, Clifford GD, et al. Rationale and design of a multidomain lifestyle program for mild cognitive impairment. Alzheimer’s & Dementia. 2026;22(1):e71068. Available from: https://alz-journals.onlinelibrary.wiley.com/doi/abs/10.1002/alz.71068. arXiv:https://alz-journals.onlinelibrary.wiley.com/doi/pdf/10.1002/alz.71068. 10.1002/alz.71068.

45. Suresha PB, Hegde C, Jiang Z, Clifford GD. An Edge Computing and Ambient Data Capture System for Clinical and Home Environments. Sensors. 2022 Jan;22(7):2511. doi:10.3390/s22072511.

46. Saghafi S, Kiarashi Y, Rodriguez AD, Levey AI, Kwon H, Clifford GD. Indoor Localization Using Multi-Bluetooth Beacon Deployment in a Sparse Edge Computing Environment. Digital Twins and Applications. 2025;2(1):e70001. doi:10.1049/dgt2.70001.

47. Julayanont P, Brousseau M, Chertkow H, Phillips N, Nasreddine ZS. Montreal Cognitive Assessment Memory Index Score (MoCA-MIS) as a Predictor of Conversion from Mild Cognitive Impairment to Alzheimer’s Disease. Journal of the American Geriatrics Society. 2014;62(4):679–84. doi:10.1111/jgs.12742.

48. King L, Horak F. On the Mini-Bestest: Scoring and the Reporting of Total Scores. Physical Therapy. 2013 Apr;93(4):571–5. arXiv:https://academic.oup.com/ptj/article-pdf/93/4/571/31640691/ptj0571.pdf. doi:10.2522/ptj.2013.93.4.571.

49. Zijlstra W, Hof AL. Assessment of spatio-temporal gait parameters from trunk accelerations during human walking. Gait & Posture. 2003;18(2):1–10. Available from: https://www.sciencedirect.com/science/article/pii/S096663620200190X. doi: 10.1016/S0966-6362(02)00190-X.

50. Christ M, Kempa-Liehr AW, Feindt M. Time Series FeatuRe Extraction on basis of Scalable Hypothesis tests (tsfresh – A Python package). Neurocomputing. 2018;307:72–7. doi:10.1016/j.neucom.2018.03.067.

51. Chzhen E, Denis C, Hebiri M, Oneto L, Pontil M. Fair Regression with Wasserstein Barycenters; 2020. Available from: https://arxiv.org/abs/2006.07286. arXiv:2006.07286.

52. Hollman JH, McDade EM, Petersen RC. Normative spatiotemporal gait parameters in older adults. Gait & Posture. 2011;34(1):111–8. Available from: https://www.sciencedirect.com/science/article/pii/S0966636211001019. doi: 10.1016/j.gaitpost.2011.03.024.

53. Bouten CVC, Koekkoek KTM, Verduin M, Kodde R, Janssen JD. A Triaxial Accelerometer and Portable Data Processing Unit for the Assessment of Daily Physical Activity. IEEE Transactions on Biomedical Engineering. 1997 Mar;44(3):136–47. doi:10.1109/10.554760.

54. Robertson DGE, Dowling JJ. Design and responses of Butterworth and critically damped digital filters. Journal of Electromyography and Kinesiology. 2003;13(6):569–73. Available from: https://www.sciencedirect.com/science/article/pii/S1050641103000804. 10.1016/S1050-6411(03)00080-4.

55. Zhang J, Veltink PH, van Asseldonk EHF. Optimizing Cut-Off Frequencies and Filter Orders for Dynamic Local Reference Frames for Human Gait Analysis in Straight-Line and Turning Tasks. In: 2024 IEEE International Symposium on Medical Measurements and Applications (MeMeA); 2024. p. 1–6. doi:10.1109/MeMeA60663.2024.10596724.

56. Virtanen P, Gommers R, Oliphant TE, Haberland M, Reddy T, Cournapeau D, et al. SciPy 1.0: fundamental algorithms for scientific computing in Python. Nature Methods. 2020 Feb;17(3):261–272. Available from: doi:10.1038/s41592-019-0686-2.

57. Del Din S, Godfrey A, Rochester L. Validation of an Accelerometer to Quantify a Comprehensive Battery of Gait Characteristics in Healthy Older Adults and Parkinson’s Disease: Toward Clinical and at Home Use. IEEE Journal of Biomedical and Health Informatics. 2016 May;20(3):838–47. doi:10.1109/JBHI.2015.2419317.

58. Patterson KK, Gage WH, Brooks D, Black SE, McIlroy WE. Evaluation of gait symmetry after stroke: A comparison of current methods and recommendations for standardization. Gait & Posture. 2010;31(2):241–6. Available from: https://www.sciencedirect.com/science/article/pii/S0966636209006493. 10.1016/j.gaitpost.2009.10.014.

59. Winter DA. Biomechanics and Motor Control of Human Movement. 4th ed. Hoboken, NJ: John Wiley & Sons; 2009.

60. Fryar CD, Gu Q, Afful J, Carroll MD, Ogden CL. Anthropometric Reference Data for Children and Adults: United States, August 2021–August 2023. National Center for Health Statistics; 2025. Available from: https://stacks.cdc.gov/view/cdc/174595.

61. Brodie MAD, Menz HB, Lord SR. Age-associated changes in head jerk while walking reveal altered dynamic stability in older people. Experimental Brain Research. 2014;232(1):51–60. doi:10.1007/s00221-013-3719-6.

62. Fulcher BD, Little MA, Jones NS. Highly comparative time-series analysis: the empirical structure of time series and their methods. Journal of The Royal Society Interface. 2013 Jun;10(83):20130048. Available from: doi:10.1098/rsif.2013.0048.

63. Rousseeuw P, Hubert M. Robust statistics for outlier detection. Wiley Interdisciplinary Reviews: Data Mining and Knowledge Discovery. 2011 01;1:73–9. doi:10.1002/widm.2.

64. Hammerla NY, Kirkham R, Andras P, Ploetz T. On preserving statistical characteristics of accelerometry data using their empirical cumulative distribution. In: Proceedings of the 2013 International Symposium on Wearable Computers. ISWC’13. New York, NY, USA: Association for Computing Machinery; 2013. p. 65–68. Available from: doi:10.1145/2493988.2494353.

65. Welch P. The use of fast Fourier transform for the estimation of power spectra: A method based on time averaging over short, modified periodograms. IEEE Transactions on Audio and Electroacoustics. 1967;15(2):70–3. doi:10.1109/TAU.1967.1161901.

66. Breiman L. Random Forests. Machine Learning. 2001;45(1):5–32. Available from: 10.1023/A:1010933404324.

67. Prokhorenkova L, Gusev G, Vorobev A, Dorogush AV, Gulin A. CatBoost: unbiased boosting with categorical features. In: Proceedings of the 32nd International Conference on Neural Information Processing Systems. NIPS’18. Red Hook, NY, USA: Curran Associates Inc.; 2018. p. 6639–6649.

68. Chen T, Guestrin C. XGBoost: A Scalable Tree Boosting System. In: Proceedings of the 22nd ACM SIGKDD International Conference on Knowledge Discovery and Data Mining. KDD’16. New York, NY, USA: Association for Computing Machinery; 2016. p. 785–794. Available from: doi:10.1145/2939672.2939785.

69. Akiba T, Sano S, Yanase T, Ohta T, Koyama M. Optuna: A Next-generation Hyperparameter Optimization Framework. In: Proceedings of the 25th ACM SIGKDD International Conference on Knowledge Discovery and Data Mining; 2019.

70. Lindvall E, Abzhandadze T, Quinn TJ, Sunnerhagen KS, Lundström E. Is the difference real, is the difference relevant: the minimal detectable and clinically important changes in the Montreal Cognitive Assessment. Cereb Circ Cogn Behav. 2024;6:100222. doi:10.1016/j.cccb.2024.100222.

71. Tamura S, Miyata K, Hasegawa S, Kobayashi S, Shioura K, Usuda S. Pooled Minimal Clinically Important Differences of the Mini-Balance Evaluation Systems Test in Patients With Early Subacute Stroke: A Multicenter Prospective Observational Study. Phys Ther. 2024;104. doi:10.1093/ptj/pzae017.

72. Bland JM, Altman D. STATISTICAL METHODS FOR ASSESSING AGREEMENT BETWEEN TWO METHODS OF CLINICAL MEASUREMENT. The Lancet. 1986 Feb;327(8476):307–10. doi:10.1016/S0140-6736(86)90837-8.

73. Ahmad MA, Eckert C, Teredesai A. Interpretable Machine Learning in Healthcare. In: Proceedings of the 2018 ACM International Conference on Bioinformatics, Computational Biology, and Health Informatics. BCB’18. New York, NY, USA: Association for Computing Machinery; 2018. p. 559–560. Available from: doi:10.1145/3233547.3233667.

74. Lundberg S, Lee SI. A Unified Approach to Interpreting Model Predictions; 2017. Available from: https://arxiv.org/abs/1705.07874. arXiv:1705.07874.

75. Dwork C, Hardt M, Pitassi T, Reingold O, Zemel R. Fairness Through Awareness;. Available from: http://arxiv.org/abs/1104.3913. arXiv:1104.3913. doi:10.48550/arXiv.1104.3913.

76. Montero-Odasso M, Verghese J, Beauchet O, Hausdorff JM. Gait and Cognition: A Complementary Approach to Understanding Brain Function and the Risk of Falling. Journal of the American Geriatrics Society. 2012 Nov;60(11):2127–36. doi:10.1111/j.1532-5415.2012.04209.x.

77. Yogev-Seligmann G, Hausdorff JM, Giladi N. The role of executive function and attention in gait. Movement Disorders. 2008;23(3):329–42. Available from: https://movementdisorders.onlinelibrary.wiley.com/doi/abs/10.1002/mds.21720. arXiv:https://movementdisorders.onlinelibrary.wiley.com/doi/pdf/10.1002/mds.21720. 10.1002/mds.21720.

78. Kourtis LC, Regele OB, Wright JM, Jones GB. Digital biomarkers for Alzheimer’s disease: the mobile/wearable devices opportunity. npj Digital Medicine. 2019;2(1):9. Available from: doi:10.1038/s41746-019-0084-2.

79. Kuan YC, Huang LK, Wang YH, Hu CJ, Tseng IJ, Chen HC, et al. Balance and Gait Performance in Older Adults with Early-Stage Cognitive Impairment. European Journal of Physical and Rehabilitation Medicine. 2021 Aug;57(4):560–7. doi:10.23736/S1973-9087.20.06550-8.

80. Fan LJ, Wang FY, Zhao JH, Zhang JJ, Li YA, Tang J, et al. From Physical Activity Patterns to Cognitive Status: Development and Validation of Novel Digital Biomarkers for Cognitive Assessment in Older Adults. International Journal of Behavioral Nutrition and Physical Activity. 2025 Jan;22(1):11. doi:10.1186/s12966-025-01706-x.

81. Soumma SB, Mamun A, Ghasemzadeh H. AI-powered Wearable Sensors for Health Monitoring and Clinical Decision Making. Current Opinion in Biomedical Engineering. 2025 Dec;36:100628. doi:10.1016/j.cobme.2025.100628.

82. Shajari S, Kuruvinashetti K, Komeili A, Sundararaj U. The Emergence of AI-Based Wearable Sensors for Digital Health Technology: A Review. Sensors. 2023 Jan;23(23):9498. doi:10.3390/s23239498.

83. Balls-Berry JJE, Babulal GM. Health Disparities in Dementia. Continuum (Minneapolis, Minn). 2022 Jun;28(3):872–84. doi:10.1212/CON.0000000000001088.

